# A six-month periodic fasting reduces microalbuminuria and improves metabolic control in patients with type 2 diabetes and diabetic nephropathy: a randomized controlled study

**DOI:** 10.1101/2021.12.01.21266958

**Authors:** Alba Sulaj, Stefan Kopf, Ekaterina von Rauchhaupt, Elisabeth Kliemank, Maik Brune, Zoltan Kender, Hannelore Bartl, Fabiola Garcia Cortizo, Katarina Klepac, Zhe Han, Varun Kumar, Valter Longo, Aurelio Teleman, Jürgen G. Okun, Jakob Morgenstern, Thomas Fleming, Julia Szendroedi, Stephan Herzig, Peter P. Nawroth

## Abstract

**Aim:** Novel dietary interventions focused on fasting, have gained scientific and public attention. Periodic fasting has emerged as a dietary modification promoting beneficial effects on metabolic syndrome. This study aimed to assess whether periodic fasting reduces albuminuria in patients with type 2 diabetes and diabetic nephropathy and determine whether a reduction in albuminuria relates to activation of nephropathy-driven pathways.

**Methods:** Forty patients with type 2 diabetes (HbA1c 7.8±0.2% [62.1±2.3 mmol/mol]) and increased albumin-to-creatinine ratio (ACR) were randomized to fasting-mimicking diet (FMD) (n=21) or Mediterranean diet (n=19) for six months with three-month follow-up. Primary endpoint was the difference of the change in ACR from baseline to after six months between study groups. Subgroup analysis for patients with micro-versus macroalbuminuria at baseline was performed. Secondary endpoints comprised HOMA-IR, circulating markers of dicarbonyl detoxification (MG-H1, glyoxalase-1 and hydroxyacetone), lipid oxidation (acylcarnitines), DNA-damage/repair, (yH2Ax) and senescence (suPAR). Comparison was done by ANCOVA adjusted for age, sex, weight loss and baseline values of the respective outcome.

**Results:** Difference of change in ACR between FMD and control group after six months was 110.3mg/g (95% CI 99.2, 121.5mg/g; P=0.45) in all patients, -30.3mg/g (95% CI -35.7, -24.9mg/g; P≤0.05] in patients with microalbuminuria, and 434.0mg/g (95% CI 404.7, 463.4mg/g; P=0.23) in those with macroalbuminuria at baseline. FMD led to change in HOMA-IR of -3,8 (95% CI -5,6, -2.0; P≤0.05) and in suPAR of - 156.6pg/ml (95% CI -172.9, -140.4pg/ml; P≤0.05) after six months, while no change was observed in markers of dicarbonyl detoxification or DNA-damage/repair. Change in AC profile was related to patient responsiveness to ACR improvement. At follow-up only HOMA-IR reduction (−1.9 [95% CI -3.7, -0.1], P≤0.05) was sustained.

**Conclusions:** When accompanied by intensive diabetes care, FMD improves microalbuminuria, HOMA-IR and suPAR levels. Lack of changes in markers of dicarbonyl detoxification and DNA-damage/repair might explain the relapse of albuminuria at follow-up.

**Trial registration:** [German Clinical Trials Register](https://www.drks.de/drks_web/navigate.do?navigationId=start) (Deutsches Register Klinischer Studien DRKS), DRKS-ID: DRKS00014287

## Introduction

Diabetic nephropathy is the most common cause of end-stage renal disease and therapeutic options for slowing its progression are limited to control of glycemia, lipidemia and blood pressure [1]. Current therapies, in particular sodium-glucose cotransporter-2 (SGLT2) inhibitors and ACE inhibitors, exert to some degree nephroprotection [2, 3]. However, the renal benefits attributed to SGLT-2 inhibitors cannot be explained by the modest improvements in glycemic, blood pressure and lipid control [2]. Once diabetic kidney disease (DKD) is established, it is only possible to slow its progression, whereas therapies for long-lasting remission are lacking [4].

Hyperglycemia may cause a decline in renal function leading to DKD via activation of the sorbitol pathway, activation of the hexosamine pathway, and an increase in oxidative stress, eventually resulting in fibrosis [5-7]. Moreover, dicarbonyl stress and AGE have long been implicated in the development of diabetic complications and DKD [8-12]. Increased DNA-damage, persistent DNA-damage signaling as well as senescence have also been described in type 2 diabetes [13-16]. Recently, it was shown in vivo that restoring DNA-repair in experimental type 1 diabetes by overexpression of a nuclear phosphomimetic RAGE reduces DNA-damage, inflammation, and might improve renal function [17].

Interestingly, recent research has shown that periodic fasting can have protective effects by reducing oxidative damage, inflammation and supporting cellular protection [18-22]. Moreover, previous work on cardiorenal benefits of SGLT-2 inhibitors postulated that enhanced ketogenesis might account for their favorable effects due to a shift in fuel metabolism from glucose oxidation to fat and subsequent utilization of ketone bodies shifting energy metabolism to ketolysis [23-26]. Since kidney metabolism relies highly on lipolysis and ketone bodies, periodic fasting might be beneficial by activating lipolysis. It is yet unclear whether periodic fasting might reduce systemic dicarbonyl stress, DNA-damage, and markers of senescence.

The first aim of this study was to test whether a periodic fasting-mimicking diet (FMD) might lead to a reduction of ACR. Secondly, we wanted to investigate whether the reduction in ACR can relate to changes in markers of dicarbonyl detoxification, fatty acid oxidation, DNA-damage/repair, and senescence.

## Methods

### Study design and study population

This randomized controlled open study was performed at the Clinics of Endocrinology, Diabetology, Metabolism and Clinical Chemistry, at the University Hospital of Heidelberg. The ethics committee of the University Hospital of Heidelberg approved all study procedures (Ethic-Nr. S-682/2016) in compliance with national guidelines and the declaration of Helsinki. This study was registered at the German Clinical Trials Register (Deutsches Register Klinischer Studien DRKS), DRKS-ID: DRKS00014287.

Patients with type 2 diabetes with good glycemic and blood pressure control, optimized treatment according to current guidelines [27], and increased albumin-to-creatinine ratio (ACR) assessed from two consecutive spot urine samples according to KDIGO guidelines were recruited [28]. All participants gave written informed consent. Inclusion criteria were age 50-75 years, eGFR>30 ml/min/173cm^2^ from CKD-EPI formula, and BMI 23-40 kg/m^2^. Exclusion criteria included acute infection/fever, severe heart, kidney, hematological or liver diseases. Withdrawal criteria included subject`s request and lack of increase in blood and/or urine ketone bodies. Full study criteria are listed in the Supplementary Data.

All participants received one individual dietary counseling before the baseline visit according to recommendations of the American Diabetes Association [27]. Baseline measures were performed before the first diet cycle. Participants were seen at baseline and each month after the diet cycle (the sixth day after the five-day diet intervention), for a total of six diet cycles. Participants were seen at refeeding timepoints: one week after the third and after the sixth diet cycle. Adherence to the study regimen was assessed from dietary protocols and measurement of ketone bodies in blood and urine. Follow-up timepoint was three months after study completion. Study design is shown in **Figure 1A**.

**Figure 1.**
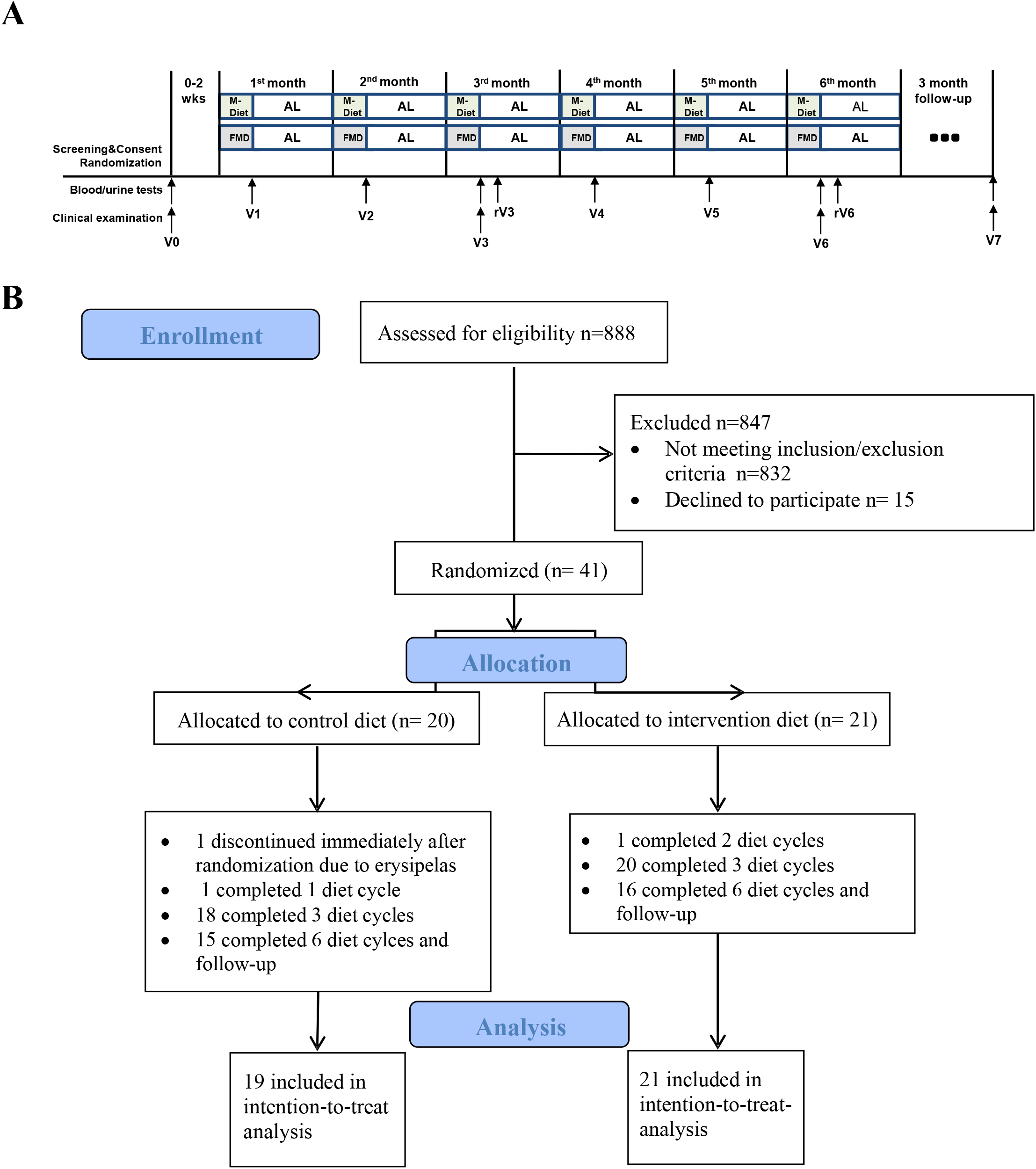
(A) Study design and (B) CONSORT flow diagram. *Abbreviations:* M-Diet Mediterranean diet, FMD fasting-mimicking diet, AL ad libitum, V0 visit at baseline, V1 visit after the first diet cycle, V2 visit after the second diet cycle, V3 visit after the third diet cycle, V4 visit after the fourth diet cycle, V5 visit after the fifth diet cycle, V6 visit after the sixth diet cycle, V7 follow-up visit three months after study completion, rV3 visit at refeeding one week after the third diet cycle and rV6 visit at refeeding one week after the sixth diet cycle.

### Randomization

Participants were randomized by a stratified computed procedure (Randomizer Version 2.0.3© Institute for Medical Informatics, Statistics and Documentation, Medical University of Graz) for BMI (cut-off value 30.0 kg/m^2^) and HbA1c (cut-off value 7.0%) to intervention or control group. The randomization list was only accessible to assigned randomization list managers.

### Diet intervention

Participants were instructed to comply for five consecutive days each month, either with FMD (intervention group) or with a Mediterranean Diet (M-Diet) (control group) and to return to their normal diet until the next diet cycle that was initiated about 25 days later. FMD was a plant-based diet that mimics fasting-like effects on glucose and ketone bodies: day 1 supplied 4600 kJ (11% protein, 46% fat, and 43% carbohydrates), whereas days 2 to 5 provided 3000 kJ (9% protein, 44% fat, and 47% carbohydrate) per day [29]. M-Diet had no change in caloric intake compared to participant’s diet before the study and was adapted according to the Mediterranean

Diet Score (MDS) [30]. Full composition of diets is shown in Supplementary Table S1. Weight loss of <10% was accepted for both study groups. Patients that were on insulin therapy were instructed to discontinue short-acting insulin and to reduce the long-acting insulin by 50% when taking FMD. Oral antidiabetic therapy was also discontinued during FMD. Glycemic levels were self-monitored (fasting and 2-hour postprandial levels, at least 4 measurements daily) with a capillary blood glucose monitoring system (Accu-Check Guide^®^, Roche) and a 24-hour telephone platform was available for participants to report hypo- or hyperglycemic episodes. Antihyperglycemic medication was reduced in case of a reduction of fasting plasma glucose by more than 20% compared to the previous measurement [29]. Antihypertensive medication was reduced if a participant reported hypotension (lower than 100 mmHg for systolic and lower than 60 mmHg for diastolic values). The aim was to maintain comparable glycemic and blood pressure control between the study groups throughout the study. All participants were instructed to avoid excessive physical activity during FMD or M-Diet and to return to their normal physical activity afterwards. Participants were asked to write their individual diet and physical activity protocol during the study. Health-related quality of life and physical health was assessed using the 12-item short-form health survey (SF-12 questionnaire) [31]. Somatic and depression symptoms were evaluated using the Patient Health Questionnaire (PHQ) [32].

### Blood Chemistry

Blood samples were drawn in fasting state and immediately processed in the Central Laboratory of the University Hospital of Heidelberg under standardized conditions. Beta-hydroxybutyrate was measured at each visit in venous blood (StatStrip® Glucose/Ketone Meter system, Nova® biomedical). Quantification of acylcarnitines (AC), methylglyoxal (MG), methylglyoxal-derived hydroimidazolone 1 (MG-H1), hydroxyacetone, glyoxalase-1 (Glo-1) activity, phosphorylated Glo-1 (pGlo-1), isolation of white and red blood cells (WBC, RBC), determination of comet tail length, yH2Ax-positivity in WBC, comet -assay and yH2Ax-positivity-measurements are shown in the Supplementary Data.

### Outcomes

Primary endpoint was defined as absolute difference of change of ACR from baseline to after six diet cycles between FMD and M-Diet group. A subgroup analysis for participants with micro-versus macroalbuminuria levels at baseline was performed for the primary endpoint. Secondary endpoints comprised the absolute difference of change of ACR after three diet cycles and the absolute difference of change after three and after six diet cycles of HOMA-IR, plasma MG-H1, plasma MG, Glo-1 activity and expression of pGlo-1 in WBC, hydroxyacetone in RBC as markers of dicarbonyl detoxification, plasma AC profile as marker of fatty acid oxidation, yH2Ax expression in WBC as marker of DNA-damage/repair and soluble urokinase plasminogen activator receptor (suPAR), as marker of kidney injury and senescence. Safety was monitored by assessment of vital signs, physical examination, electrocardiogram, adverse events following the Common Terminology Criteria for Adverse Events (CTCAE; v4.0), and laboratory results at each visit.

### Power Calculation

According to previous studies, a reduction of albuminuria by at least 30% is considered clinically relevant [33, 34]. Thus, the actual study required a total sample size of n=34 patients to detect a mean difference between groups of 30% decrease of ACR from baseline, assuming a standard deviation of 30% for both groups with a two-sample t-test and a two-sided significance level of α=5% and a power of at least 80%. An estimated dropout rate of 15% resulted in 40 participants. Sample size calculation was performed in PASS 16.0.12.

### Statistical Analysis

Analyses were performed in the intention-to-treat population, of which at least the baseline and three and/or sixth months data were available. Missing values were not imputed. Available-case analysis was performed, and the respective sample size is reported. Patient characteristics are shown as mean±SEM for normally distributed variables, median with interquartile range (IQR) for log-normally distributed variables, and frequencies for categorical variables. Comparison of changes between FMD and M-Diet group was done by an ANCOVA adjusted for age, sex, weight loss, and the respective baseline value of the outcome. For the secondary endpoints, a hierarchical test procedure was applied to account for multiple testing. Statistical computing was performed using R (version 3.6.1 2019-07-05). Visualization of the data was performed employing R core packages, tiduverse, pheatmap and GraphPad Prism version 5.00 for Windows (GraphPad Software, San Diego California USA).

## Results

### Patient characteristics

Between March 2018 and May 2020, 41 patients were randomized to FMD group (n=21) or M-diet group (n=20). One patient of the M-Diet group discontinued immediately after randomization due to erysipelas and is not included in the analysis. 38 (95%) patients completed three diet cycles and 33 (83%) completed the study. 31 (78%) were seen at follow-up (follow-up time was 112±31 days). The main reason for withdrawal after the third diet cycle and at follow-up were the restrictions related to the Covid-19 pandemic. Study CONSORT flow diagram is shown in **Figure 1B**.

Baseline anthropometric and metabolic measures were comparable between groups (**Table 1**). Health-related physical activity assessed from SF-12 questionnaire and daily calorie intake, assessed from self-reported diet protocols, did not differ at baseline, and during the study there was no change between the study groups after 5 days of FMD or M-Diet (data not shown).

**Table 1.**
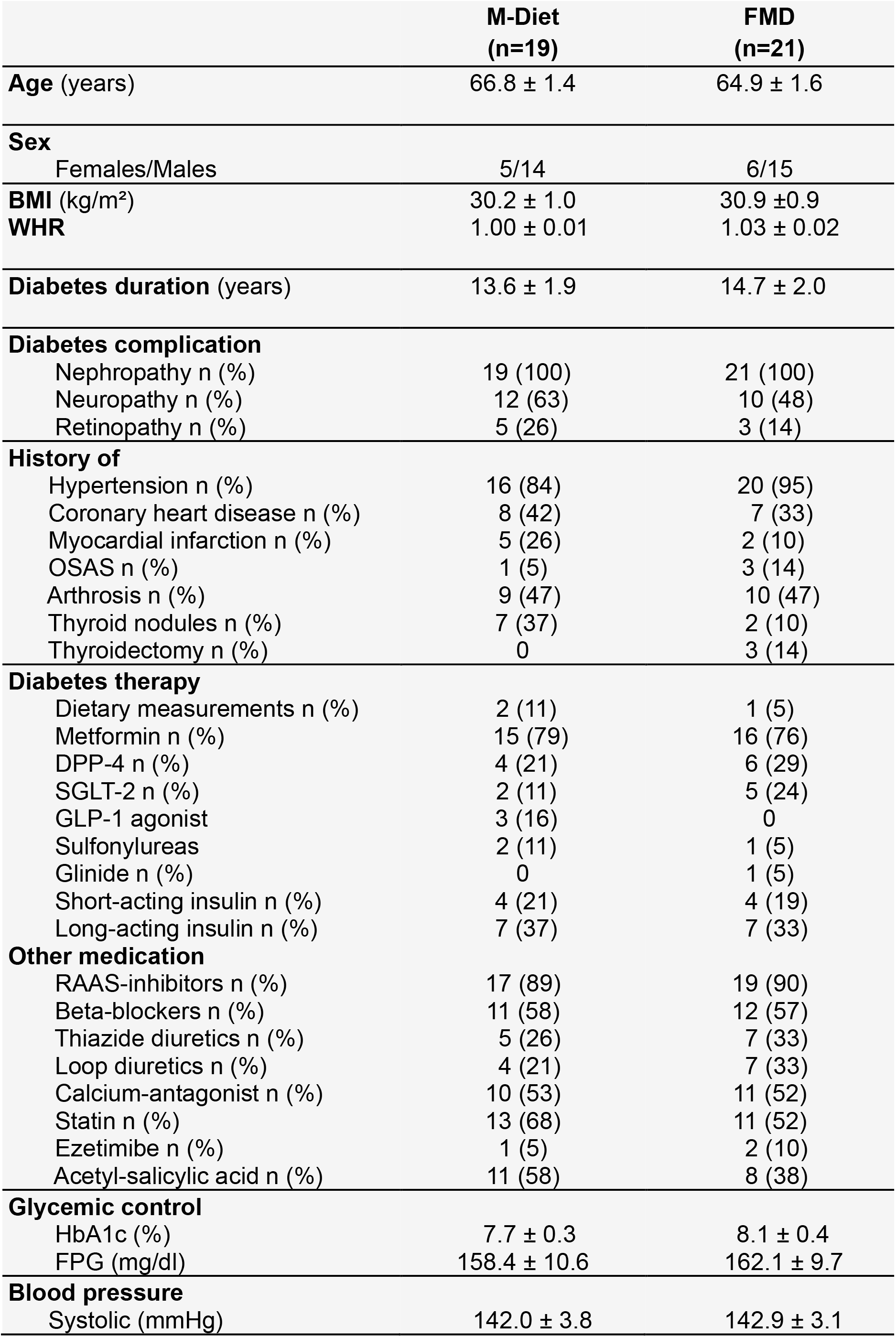
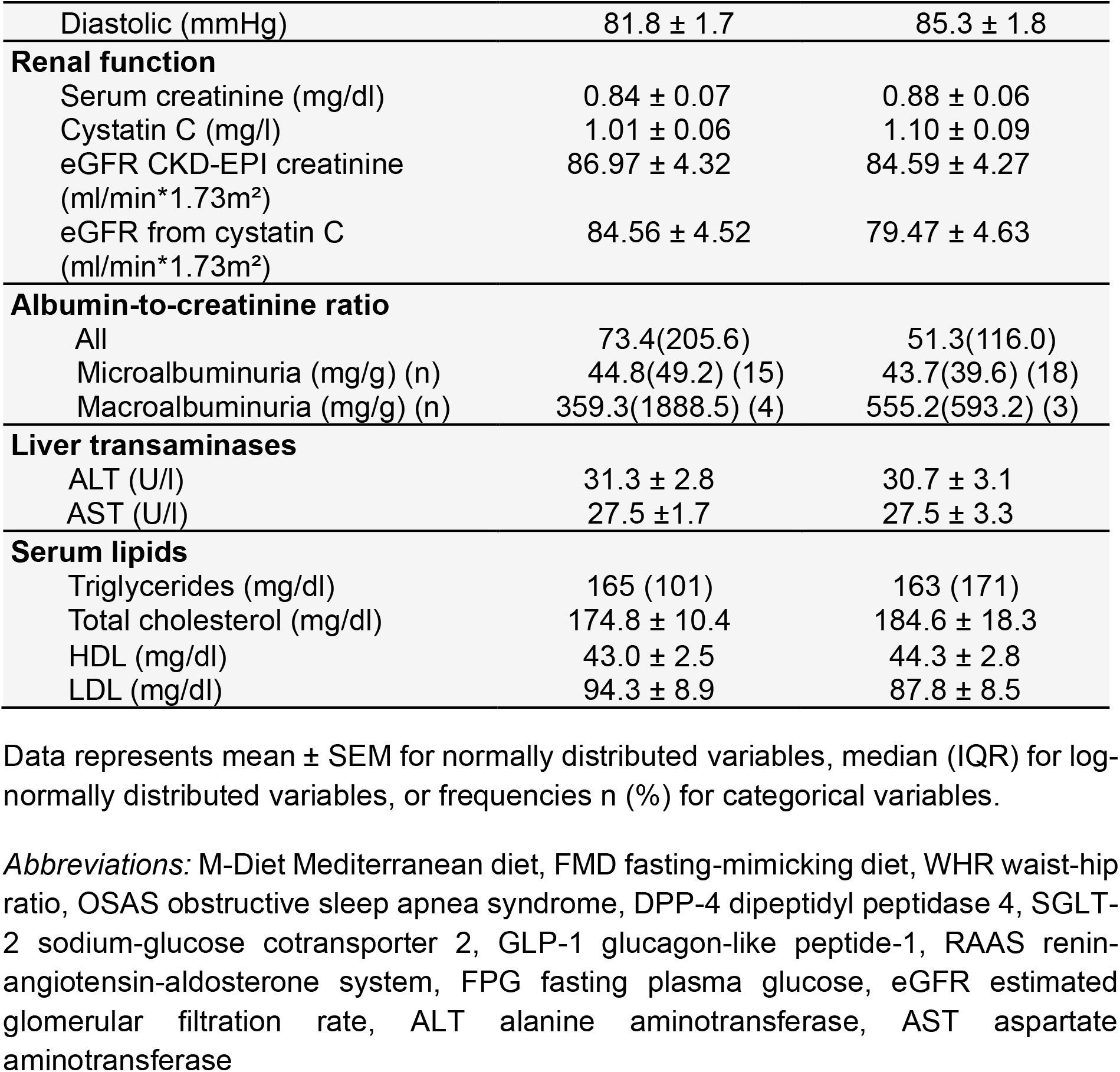
Patient characteristics at baseline.

|  | **M-Diet**<br>**(n=19)** | **FMD**<br>**(n=21)** |
| --- | --- | --- |
| **Age** (years) | 66.8 ± 1.4 | 64.9 ± 1.6 |
| **Sex** |  |  |
| Females/Males | 5/14 | 6/15 |
| **BMI** (kg/m<sup>2</sup>) | 30.2 ± 1.0 | 30.9 ± 0.9 |
| **WHR** | 1.00 ± 0.01 | 1.03 ± 0.02 |
| **Diabetes duration** (years) | 13.6 ± 1.9 | 14.7 ± 2.0 |
| **Diabetes complication** |  |  |
| Nephropathy n (%) | 19 (100) | 21 (100) |
| Neuropathy n (%) | 12 (63) | 10 (48) |
| Retinopathy n (%) | 5 (26) | 3 (14) |
| **History of** |  |  |
| Hypertension n (%) | 16 (84) | 20 (95) |
| Coronary heart disease n (%) | 8 (42) | 7 (33) |
| Myocardial infarction n (%) | 5 (26) | 2 (10) |
| OSAS n (%) | 1 (5) | 3 (14) |
| Arthrosis n (%) | 9 (47) | 10 (47) |
| Thyroid nodules n (%) | 7 (37) | 2 (10) |
| Thyroidectomy n (%) | 0 | 3 (14) |
| **Diabetes therapy** |  |  |
| Dietary measurements n (%) | 2 (11) | 1 (5) |
| Metformin n (%) | 15 (79) | 16 (76) |
| DPP-4 n (%) | 4 (21) | 6 (29) |
| SGLT-2 n (%) | 2 (11) | 5 (24) |
| GLP-1 agonist | 3 (16) | 0 |
| Sulfonylureas | 2 (11) | 1 (5) |
| Glinide n (%) | 0 | 1 (5) |
| Short-acting insulin n (%) | 4 (21) | 4 (19) |
| Long-acting insulin n (%) | 7 (37) | 7 (33) |
| **Other medication** |  |  |
| RAAS-inhibitors n (%) | 17 (89) | 19 (90) |
| Beta-blockers n (%) | 11 (58) | 12 (57) |
| Thiazide diuretics n (%) | 5 (26) | 7 (33) |
| Loop diuretics n (%) | 4 (21) | 7 (33) |
| Calcium-antagonist n (%) | 10 (53) | 11 (52) |
| Statin n (%) | 13 (68) | 11 (52) |
| Ezetimibe n (%) | 1 (5) | 2 (10) |
| Acetyl-salicylic acid n (%) | 11 (58) | 8 (38) |
| **Glycemic control** |  |  |
| HbA1c (%) | 7.7 ± 0.3 | 8.1 ± 0.4 |
| FPG (mg/dl) | 158.4 ± 10.6 | 162.1 ± 9.7 |
| **Blood pressure** |  |  |
| Systolic (mmHg) | 142.0 ± 3.8 | 142.9 ± 3.1 |
| Diastolic (mmHg) | 81.8 ± 1.7 | 85.3 ± 1.8 |
| **Renal function** |  |  |
| Serum creatinine (mg/dl) | 0.84 ± 0.07 | 0.88 ± 0.06 |
| Cystatin C (mg/l) | 1.01 ± 0.06 | 1.10 ± 0.09 |
| eGFR CKD-EPI creatinine (ml/min*1.73m<sup>2</sup>) | 86.97 ± 4.32 | 84.59 ± 4.27 |
| eGFR from cystatin C (ml/min*1.73m<sup>2</sup>) | 84.56 ± 4.52 | 79.47 ± 4.63 |
| **Albumin-to-creatinine ratio** |  |  |
| All | 73.4(205.6) | 51.3(116.0) |
| Microalbuminuria (mg/g) (n) | 44.8(49.2) (15) | 43.7(39.6) (18) |
| Macroalbuminuria (mg/g) (n) | 359.3(1888.5) (4) | 555.2(593.2) (3) |
| **Liver transaminases** |  |  |
| ALT (U/l) | 31.3 ± 2.8 | 30.7 ± 3.1 |
| AST (U/l) | 27.5 ± 1.7 | 27.5 ± 3.3 |
| **Serum lipids** |  |  |
| Triglycerides (mg/dl) | 165 (101) | 163 (171) |
| Total cholesterol (mg/dl) | 174.8 ± 10.4 | 184.6 ± 18.3 |
| HDL (mg/dl) | 43.0 ± 2.5 | 44.3 ± 2.8 |
| LDL (mg/dl) | 94.3 ± 8.9 | 87.8 ± 8.5 |

Data represents mean ± SEM for normally distributed variables, median (IQR) for log-normally distributed variables, or frequencies n (%) for categorical variables.
*Abbreviations:* M-Diet Mediterranean diet, FMD fasting-mimicking diet, WHR waist-hip ratio, OSAS obstructive sleep apnea syndrome, DPP-4 dipeptidyl peptidase 4, SGLT-2 sodium-glucose cotransporter 2, GLP-1 glucagon-like peptide-1, RAAS renin-angiotensin-aldosterone system, FPG fasting plasma glucose, eGFR estimated glomerular filtration rate, ALT alanine aminotransferase, AST aspartate aminotransferase

### Effect of FMD on albuminuria and risk factors for DKD

Difference of change in ACR between FMD and M-Diet group after six diet cycles was 110.3mg/g (99.2, 121.5mg/g; P=0.45) in all patients (**Table 2**). In the subgroup analysis, the difference of change in ACR between groups after six diet cycles was - 30.3mg/g (−35.7, -24.9mg/g; P≤0.05] in patients with microalbuminuria, corresponding to a 54% reduction (**Figure 2A** and **Table 2**), and 434.0mg/g (404.7, 463.4mg/g; P=0.23) in those with macroalbuminuria (**Table 2**). Difference of change in ACR between groups after three diet cycles was -62.5mg/g (−71.3, -53.7mg/g; P=0,45) in all patients (**Table 2**), -23.7mg/g (−28.4, -18.9 mg/g; P≤0.05) in patients with microalbuminuria, corresponding to a 37% reduction (**Figure 2A** and **Table 2**), and - 176.2mg/g (−203.1, -149.4mg/g; P=0.47) in patients with macroalbuminuria (**Table 2**). FMD resulted in a reduction of urea levels after three (−11.8mg/dl [-14.0, -9.7mg/dl]; P≤0.01) and six diet cycles (−17.9mg/dl [-21.2, -14.6mg/dl]; P≤0.05) compared to M-Diet (**Table 2**). Likewise, FMD intervention led to less decline in eGFR based on cystatin C after six diet cycles resulting in a difference of change of 8.3ml/min/1,73m^2^ (5.9, 10.6ml/min/1,73m^2^; P≤0.05) between study groups. After three diet cycles the difference of change in eGFR based on cystatin C between the groups was 2.4ml/min/1,73m^2^ (0.7, 4.1ml/min/1,73m^2^; P=0.06) (**Table 2**). suPAR decreased after three (difference of change between the groups -214.8pg/ml [-238.4, -191.3pg/ml]; P≤0.05) and after six diet cycles (difference of change between groups -156.6pg/ml [-172.9, -140.4pg/ml]; P≤0.05) (**Table 2**).

**Table 2.** Metabolic and anthropometric parameters at baseline (V0), after 3 months (V3), after 6 months (V6) and at follow-up (V7).

| Parameter | Group | V0 | V3 | V6 | V7 |
| --- | --- | --- | --- | --- | --- |
| **Glycemia** |  |  |  |  |  |
| FPG<br>(mg/dl) | M-Diet | 158.4±10.6<br>(19) | 149.1±11.4<br>(18) | 151.2±12.1<br>(16) | 167.5±18.2<br>(15) |
|  | FMD | 162.1±9.7<br>(21) | 146.7±13.8<br>(20) | 137.4±12.7<br>(17) | 151.9±8.5<br>(16) |
| **HbA1c**<br>(%) | M-Diet | 7.7±0.3(19) | 7.2±0.3(18) | 7.7±0.3(16) | 8.0±0.4(15) |
|  | FMD | 8.1±0.4(21) | 7.2±0.3(20) | **6.7±0.3(17)\*** | 7.6±0.4(16) |
| C-Peptid<br>(ng/ml) | M-Diet | 3.3±0.4(19) | 2.9±0.4(18) | 3.2±0.6(16) | 3±0.5(14) |
|  | FMD | 3.1±0.4(21) | 2.3±0.4(20) | 2.0±0.3(17) | 2.7±0.3(15) |
| **HOMA-IR** | M-Diet | 6.1±0.9(13) | 6.0±0.9 (13) | 5.7±1.0 (11) | 6.1±0.9(10) |
|  | FMD | 6.4±1.9(14) | 4.9±2.7(14) | **2.6±0.6(12)\*\*** | **5.1±1.7(10)\*** |
| **Blood pressure** |  |  |  |  |  |
| Systolic BP | M-Diet | 142.0±3.8<br>(19) | 144.6±5.9<br>(18) | 142.5±4.1<br>(16) | 136.6±2.9<br>(14) |
|  | FMD | 142.9±3.1<br>(21) | 135.9±3.2<br>(19) | 141.3±3.2<br>(16) | 142.6±5.3<br>(14) |
| Diastolic BP | M-Diet | 81.8±1.7(19) | 81.3±2.1(18) | 81.8±2.4(16) | 80.5±2.3(14) |
|  | FMD | 85.3±1.8(21) | 81.6±1.5(19) | 83.2±1.8(16) | 85.6±2.3(14) |
| **Lipidemia** |  |  |  |  |  |
| Cholesterol<br>(mg/dl) | M-Diet | 174.8±10.4<br>(19) | 171.7±9.9<br>(18) | 184.2±11.2<br>(16) | 188.7±13.4<br>(15) |
|  | FMD | 184.6±18.3<br>(21) | 164.7±13.1<br>(20) | 160.2±10.3<br>(17) | 177.8±11.9<br>(16) |
| LDL<br>(mg/dl) | M-Diet | 94.3±8.9<br>(18) | 90.2±7.5<br>(18) | 96.6±8.4<br>(16) | 103.8±12.3<br>(13) |
|  | FMD | 87.8±8.5<br>(20) | 95.2±11.5<br>(19) | 87.2±9.6<br>(17) | 84.6±10.0<br>(14) |
| HDL<br>(mg/dl) | M-Diet | 43.0±2.5(19) | 44.3±2.7(18) | 46.8±2.6(16) | 45.7±3.7(15) |
|  | FMD | 44.3±2.8(21) | 43.3±2.6(20) | 48.8±3.3(17) | 51.7±3.8(16) |
| TG<br>(mg/dl) | M-Diet | 205.6±25.8<br>(19) | 185.5±20.5<br>(18) | 204.8±24.6<br>(16) | 214.7±35.2<br>(15) |
|  | FMD | 277.2±75.6<br>(21) | 166.8±37.5<br>(20) | 121.2±12.4<br>(17) | 209.4±35.8<br>(16) |
| Lp(a)<br>(mg/dl) | M-Diet | 30.3±8.7<br>(19) | 35.6±10.5<br>(17) | 33.6±9.7<br>(16) | 34.6±10.7<br>(14) |
|  | FMD | 16.3±3.5(21) | 20.6±4.4(20) | 17.5±2.1(17) | 14.3±1.7(15) |
| **Liver parameters** |  |  |  |  |  |
| AST<br>(U/l) | M-Diet | 27.5±1.7(19) | 31.1±3.0(18) | 27.2±1.6(16) | 26.3±1.5(15) |
|  | FMD | 27.5±3.3(21) | 29.9±2.4(20) | 25.5±1.4(17) | 23.0±1.6(16) |
| ALT<br>(U/l) | M-Diet | 31.3±2.8(19) | 32.8±4.4(18) | 30.4±2.8(16) | 30.7±2.7(15) |
|  | FMD | 30.6±3.1(21) | 34.7±3.5(20) | 25.6±1.9(17) | 23.3±2.4(16) |
| Liver<br>stiffness(kPa) | M-Diet | 6.9±0.8(19) | 6.4±0.6(18) | 6.0 ±0.6(16) | 7.2±1.2(14) |
|  | FMD | 8.1±1(21) | 6.2±0.6(19) | 6.4±0.7(17) | 7.1±0.8(15) |
| **Protein Level** |  |  |  |  |  |
| Plasma-Albumin<br>(g/l) | M-Diet | 44.3±0.7(19) | 44.8±0.6(18) | 44.9±0.7(16) | 44.6±0.7(15) |
|  | FMD | 45.9±0.4(21) | 46.7±0.5(20) | 47.3±0.6(17) | 45.5±0.5(16) |
| Plasma-total<br>protein (g/l) | M-Diet | 72.9±1.4(19) | 72.8±1.4(18) | 72.4±1.5(16) | 71.5±1.4(15) |
|  | FMD | 73.3±0.9(21) | 74.6±1.1(20) | 72.9±0.9(17) | 69.5±0.8(16) |

| Renal parameters |  |  |  |  |  |
| --- | --- | --- | --- | --- | --- |
| uACR<br>(mg/g) | M-Diet | 73.4(205.6)<br>(19) | 72.7(125.8)<br>(18) | 63.1(353.6)<br>(16) | 76.0(149.9)<br>(15) |
|  | FMD | 51.3(116.0)<br>(21) | 34.8(43.6)<br>(20) | 25.6(40.3)<br>(17) | 36.5(137.8)<br>(16) |
| **Micro.uACR**<br>**(mg/g)** | M-Diet | 44.8(49.2)<br>(14) | 35.8(58.0)<br>(13) | 37.2(50.5)<br>(11) | 22.9(65.6) (11) |
|  | FMD | 43.7(39.6)<br>(17) | **29.1(36.1)**<br>**(16)\*** | **23.4(30.3)**<br>**(14)\*** | 27.4(27.1) (13) |
| Macro.uACR<br>(mg/g) | M-Diet | 359.3(1888.5)<br>(4) | 249.2(2533.6)<br>(4) | 430.3(592.5)<br>(3) | 399.8(143.4)<br>(3) |
|  | FMD | 555.2(593.2)<br>(3) | 666.1(575.0)<br>(3) | 1034.7(591.1)<br>(3) | 915.4(905.8)<br>(3) |
| **Urea**<br>**(mg/dl)** | M-Diet | 39.1±3.2(19) | 38.1±2.9(18) | 48.8±8.7(16) | 35.9±2.3(15) |
|  | FMD | 38.8±4(21) | **25.6±1.9(20)\*\*** | **26.2±2.1(17)\*** | 38.2±2.9(16) |
| Creatinine<br>(mg/dl) | M-Diet | 0.8±0.1(19) | 0.9±0.1(18) | 0.8±0.1(16) | 0.8±0.1(15) |
|  | FMD | 0.9±0.1(21) | 0.9±0.1(20) | 0.9±0.1(17) | 0.9±0.1(16) |
| eGFR CKD-EPI(creatinine)<br>(ml/min/1.73m<sup>2</sup>) | M-Diet | 87±4.3(19) | 85±4.5(18) | 86.4±4.7(16) | 87.7±4.3(15) |
|  | FMD | 84.6±4.3(21) | 83.2±4.2(20) | 81.8±4.8(17) | 82.3±4.7(16) |
| **Cystatin C**<br>**(mg/l)** | M-Diet | 1±0.1(19) | 1.1±0.1(18) | 1.2±0.1(16) | 1.1±0.1(15) |
|  | FMD | 1.1±0.1(21) | 1.1±0.1(20) | **1.1±0.1(17)\*** | 1.1±0.1(16) |
| **eGFR (Cystatin**<br>**C)(ml/min/1.73m<sup>2</sup>)** | M-Diet | 84.6±4.5(19) | 80±4.3(18) | 72.9±5.6(16) | 73.3±4.9(15) |
|  | FMD | 79.5±4.6(21) | 77.2±4.5(20) | **77.4±5.1(17)\*** | 71.2±4.9(16) |
| **sUPAR**<br>**(pg/ml)** | M-Diet | 3040.5±218.0<br>(19) | 3106.0±335.6<br>(17) | 2975.4±187.6<br>(16) | 3052.8±197.0<br>(14) |
|  | FMD | 3122±375<br>(21) | **2998.6±216.9**<br>**(20)\*** | **2737.1±117.8**<br>**(17)\*** | 3036.7±180.4<br>(15) |
| Ketogenesis |  |  |  |  |  |
| **Blood ketones**<br>**(mmol/l)** | M-Diet | 0.1±0.0(19) | 0.2±0.0(18) | 0.1±0.0(16) | 0.1±0.0(14) |
|  | FMD | 0.2±0.0(21) | **0.5±0.1(20)\*\*** | **0.5±0.1(17)\*\*** | 0.1±0.0(15) |
| Urine ketones<br>(mg/dl) | M-Diet | 0.3±0.3(19) | 2.2±0.9(18) | 2.5±1.3(16) | 1.8±1.1(14) |
|  | FMD | 1.2±0.8(21) | 5.8±1.5(20) | 7.9±3.2(17) | 1.0±0.5(15) |
| Other parameters |  |  |  |  |  |
| Uric acid<br>(mg/dl) | M-Diet | 6.0±0.3(19) | 6.1±0.3(18) | 6.0±0.4(16) | 5.9±0.3(14) |
|  | FMD | 6.6±0.4(21) | 6.4±0.3(20) | 6.8±0.4(17) | 6.4±0.5(15) |
| hsCRP<br>(mg/l) | M-Diet | 1.9±0.4(19) | 2.1±0.5(18) | 2.2±0.6(16) | 3.2±1.0(15) |
|  | FMD | 5.1±1.6(21) | 3.2±1.0(20) | 2.6±1.2(17) | 2.3±0.9(16) |
| hsTNT<br>(pg/ml) | M-Diet | 15.9±2.2(19) | 17.0±2.5(18) | 18.3±3.6(16) | 17.1±3.1(15) |
|  | FMD | 12.6±1.5(21) | 12.7±1.6(20) | 13.0±1.3(17) | 15.9±2.7(16) |
| IGF-1<br>(ng/ml) | M-Diet | 111.9±8.5<br>(19) | 117.2±9.8<br>(18) | 115.1±10.3<br>(16) | 123.7±12.6<br>(14) |
|  | FMD | 135.1±8.7<br>(21) | 127.8±7.0<br>(20) | 127.9±9.1<br>(16) | 137.2±7.7<br>(15) |
| IL-6<br>(pg/ml) | M-Diet | 3.0±0.6(19) | 3.8±0.6(18) | 3.2±0.8(15) | 2.9±0.4(14) |
|  | FMD | 2.9±0.4(21) | 3.5±0.6(20) | 2.3±0.1(17) | 2.3±0.2(14) |
| NT-proBNP<br>(ng/l) | M-Diet | 149.7±39.1<br>(19) | 169.2±34.1<br>(17) | 129.0±32.2<br>(16) | 140.4±41.4<br>(15) |
|  | FMD | 198.5±100.3<br>(21) | 202.2±70.0<br>(19) | 173.5±57.4<br>(17) | 252.8±111.2<br>(16) |
| **Plasma MG-H1**<br>**nM** | M-Diet | 217.9±36.0<br>(19) | 132.7±15.4<br>(18) | 246.0±27.9<br>(16) | 281.1±37.6<br>(15) |
|  | FMD | 213.3±32.0<br>(21) | **81.7±10.7**<br>**(20)\*\*\*** | 297.4±72.0<br>(17) | 294.7±50.31<br>(15) |

| Body weight |  |  |  |  |  |
| --- | --- | --- | --- | --- | --- |
| Body weight (kg) | M-Diet | 93.3±3.6(19) | 93.0±3.9(18) | 93.1±4.4(16) | 90.5±4.5(14) |
|  | FMD | 95.7±3.6(21) | **90±3.8(20)\*\*\*** | **85.8±3.6(17)\*\*\*** | 91.4±3.7(15) |
| BMI (kg/m<sup>2</sup>) | M-Diet | 30.2±1.0(19) | 30.1±1.1(18) | 30.2±1.2(16) | 29.8±1.3(14) |
|  | FMD | 31±0.9(21) | **29.1±1.1(20)\*\*\*** | **27.7±1.0(17)\*\*\*** | 29.4±1.1(15) |
| Body composition (BIA) |  |  |  |  |  |
| Fat mass (%) | M-Diet | 29.9±1.9(18) | 30.1±1.9(17) | 30.2±2.1(15) | 30.3±2.2(14) |
|  | FMD | 31.9±1.8(21) | 31.1±1.9(20) | 29.6±2.0(16) | 30.1±2.2(15) |
| Fat mass (kg) | M-Diet | 28.4±2.7(18) | 28.4±2.8(17) | 28.5±2.9(15) | 27.9±3.0(14) |
|  | FMD | 30.7±2.1(21) | 28.4±2.4(20) | 25.3±2.2(16) | 27.7±2.4(15) |
| Fat free mass (%) | M-Diet | 70.1±1.9(18) | 69.9±1.9(17) | 69.7±2.1(15) | 70.4±2.6(14) |
|  | FMD | 68.1±1.8(21) | 68.9±1.9(20) | 70.5±2.1(16) | 69.9±2.2(15) |
| Fatt free mass (kg) | M-Diet | 64.8±2.4(18) | 63.8±2.4(17) | 63.7±2.8(15) | 62.7±3.0(14) |
|  | FMD | 64.9±2.7(21) | 61.5±2.6(20) | 59.4±2.7(16) | 63.7±3.1(15) |
| Body liquid (%) | M-Diet | 53.2±1.6(18) | 52.9±1.5(17) | 52.5±1.6(15) | 52.4±1.8(14) |
|  | FMD | 51±1.4(21) | 51.2±1.4(20) | 51.7±1.5(16) | 52.7±1.5(15) |
| Body liquid (l) | M-Diet | 49.2±1.9(18) | 48.3±1.8(17) | 48.0±2.2(15) | 47.2±2.4(14) |
|  | FMD | 48.8±2.4(21) | 46.0±2.3(20) | 43.8±2.3(16) | 48.2±2.5(15) |
| Phase angle | M-Diet | 5.8±0.1(18) | 5.8±0.1(17) | 5.8±0.1(15) | 5.6±0.1(14) |
|  | FMD | 5.8±0.2(21) | 5.7±0.2(20) | 5.7±0.2(16) | 5.5±0.2(15) |

Data are shown as mean ± SEM of unadjusted values of variable for normally distributed variables or as median (IQR) for log-normally distributed variables. Statistically significant values are written in bold and indicate significance level of intervention effect on change of parameter compared to baseline and are based on M-Diet corrected ANCOVA with adjustment for age, sex, and weight loss. \*\*\* P≤0.001, \*\*P≤0.01, \*P≤0.05.
*Abbreviations:* M-Diet Mediterranean diet, FMD fasting-mimicking diet, FPG fasting plasma glucose, HOMA-IR Homeostatic Model Assessment of Insulin Resistance, TG triglyceride, Lp(a) lipoprotein (a), ALT alanine aminotransferase, AST aspartate aminotransferase, uACR urinary albumin-to-creatinine ratio, Micro.uACR microalbuminuria, Macro.uACR macroalbuminuria, eGFR estimated glomerular filtration rate, sUPAR soluble urokinase-type plasminogen activator receptor, hsCRP high-sensitivity C-reactive protein, hsTNT high-sensitivity troponin T, IL-6 interleukin 6, NT-proBNP N-terminal prohormone of brain natriuretic peptide, BIA bioimpedance analysis.

**Figure 2.**
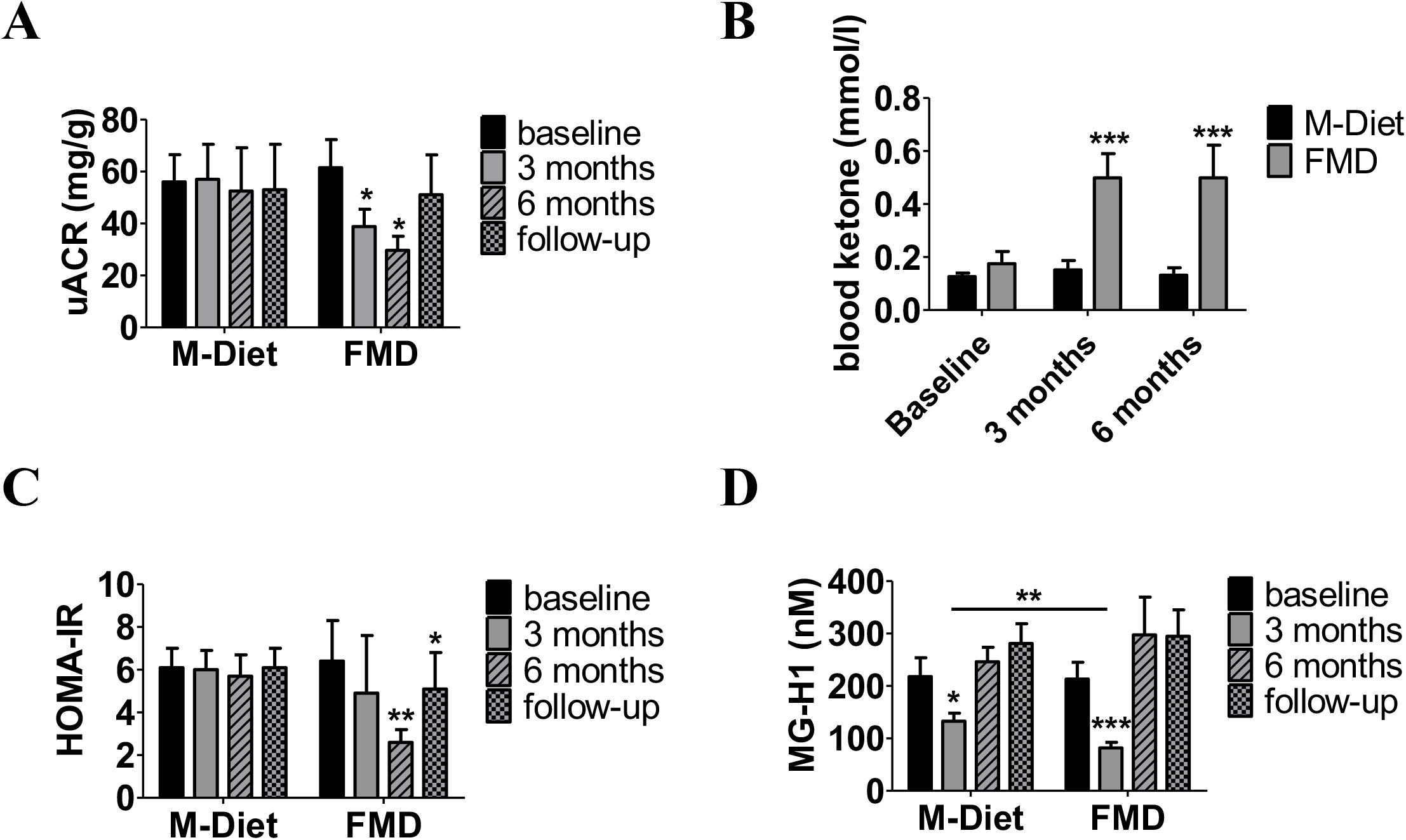
Effects of fasting on microalbuminuria (A), ketone bodies (B), HOMA-IR (C) and MG-H1 (D), after 3 months, after 6 months and at follow-up. Data are shown as mean ± SEM of unadjusted values of parameter. *P*-values indicate significance level of intervention effect on change compared to baseline and are based on M-Diet corrected ANCOVA with adjustment for age, sex and weight loss. *** P≤0.001, **P≤0.01, *P≤0.05. *Abbreviations:* M-Diet Mediterranean diet, FMD fasting-mimicking diet, uACR urinary albumin-to-creatinine ratio, HOMA-IR Homeostatic Model Assessment of Insulin Resistance, MG-H1 methylglyoxal-derived hydroimidazolone 1.

After six diet cycles FMD led to a 17 % reduction in HbA1c value compared to baseline and M-Diet group (−1.1% [-1.6, -0.6], P≤0.05, **Table 2**), while no change was observed after three diet cycles. However, fasting plasma glucose (FPG) was comparable and did not change between study groups after six diet cycles (**Table 2**). HOMA-IR decreased by 60% after 6 diet cycles (−3,8 [-5,6, -2.0]; P≤0.05) in the non-insulin treated participants of FMD group corrected to M-Diet and baseline values (**Figure 2C** and **Table 2**).

There was no change observed in LDL, HDL, or triglyceride levels in the FMD group throughout the study compared to M-diet group and baseline (**Table 2**). Weight loss of -5.6kg (−5.9%) (−6.5, -4.6, P≤0.001) after three diet cycles and of -7.2kg (−7.5%) (−9.4, -4.9, P≤0.001) after six diet cycles was observed in the FMD group, compared to a weight loss of -0.7kg (−0.8%) (−2.1, 0.8) after three diet cycles and -1.1kg (−1.2%) (−2.9, 0.7) after six diet cycles in the M-diet group. However, when adjusted for weight loss, body composition analysis revealed no change between the two study groups, neither in fat nor in fat-free mass (**Table 2**).

When exploring for protective mechanisms that could explain the reduction in ACR, we found after three diet cycles a reduction of plasma MG-H1 by 62% in the FMD group (−131.6nM [-185.2, -78.0], P≤0.001) and by 39% in the M-diet group (−92,2nM [-161.2, -23.2], P≤0.05, **Figure 2D** and **Table 2**). The MG-H1 reduction in the FMD group was higher than the reduction in the M-diet group leading to a difference of change in MG-H1 of -39,4nM (−46,7, -32,1nM; P≤0.01) between the groups. After six FMD cycles difference of change in MG-H1 was comparable to baseline (absolute change 39,5nM [-91.3, 170.3], P=0.13, **Figure 2D** and **Table 2**). The change in MG-H1 didn’t associate with the change in ACR, either in the M-Diet group or in the FMD group (data not shown). Moreover, we found no change in plasma MG, Glo-1 activity or pGlo-1 expression in WBC and no change in hydroxyacetone levels in RBC (**Supplementary Figure 3A-3D**) or yH2Ax levels in WBC (**Supplementary Figure 3E and 3F**). Both study groups were mainly comprised of male participants (74% of the M-Diet group and 71% of the FMD group), and no differences in primary or secondary endpoints were found between female and male participants (data not shown).

### Effect of FMD on fatty acid oxidation and amino acid metabolism

Participants of the FMD group who showed at least an 30% improvement in ACR (referred to as responders) elicited a distinct response in AC profile compared to the non-responders, especially in acetylcarnitine levels (C2) (**Supplementary Figure 4**). Notably, C2 levels increased after three diet cycles in responders (4.2μmol/L [0.2, 8.2], P≤0.01) compared to non-responders (−1.9μmol/L [-4.5, 0.8]) and to the M-diet group (0.7μmol/L [-0.7, 2.1] (**Supplementary Table S2**). A similar increase was observed also after six diet cycles (P≤0.01, **Supplementary Table S2**). Alanine concentration in responders after six diet cycles was reduced (−48.4μmol/L [-78.8, -18.1], P≤ 0.05), whereas glycine concentration was increased (39.6μmol/L [18.8, 60.4], P≤0.05, **Supplementary Table S2**).

When analyzing for parameters that could explain the distinct metabolic response between responders and non-responders in both study groups, we found no differences in anthropometric and metabolic parameters or body weight (**Supplementary Table S3**). Moreover, no differences in medication at baseline and throughout the study was observed between responders and non-responders (data not shown). Of note, responders of both groups showed lower ACR levels already at baseline (FMD responders 85.7mg/g [46.7, 124.7], compared to FMD non-responders 271mg/g (20, 541.2; P≤0.05) and M-Diet responders 55.5mg/g (23.3, 87.7) compared to M-Diet non-responders 563.0mg/g (6, 1120; P≤0.05) (**Supplementary Table S3**).

### Partially sustained changes at refeeding and at follow-up

At refeeding (one week after the third and sixth diet cycle), participants of both study groups had comparable HbA1c, FPG, LDL, HDL, triglyceride, HOMA-IR, eGFR, ACR, plasma MG-H1 and suPAR levels (**Supplementary Table S4**). At follow-up, only the reduction in HOMA-IR was sustained (−1.9 [-3.7, -0.1], P≤0.05, **Figure 2A** and **Table 2**), whereas no change in body weight or body composition was reported compared to baseline (**Table 2**). Apart from three participants of the FMD group, all other participants had to some degree increased the antihyperglycemic medication (data not shown). The changes reported during FMD on ACR, MG-H1 plasma levels, and on AC plasma profile were reversed to baseline values at follow-up (**Figure 2B-2D** and **Table 2**).

### Safety of FMD and effect on antihyperglycemic and antihypertensive medication

FMD was well tolerated with 71 to 95% of the participants (depending on the adverse event) reporting no adverse effects. The most common self-reported grade 1 (mild) or grade 2 (moderate) symptoms experienced were weakness, muscle pain, dizziness, and headache. No adverse effects of grade 3 or higher were reported (**Supplementary Figure 1A**). Hypoglycemic episodes experienced during diet intervention did not differ between the study groups (grade 1 hypoglycemic episodes: 10% (FMD) vs 11% (M-Diet), grade 2 hypoglycemic episodes 5% (FMD) vs 11% (M-Diet), **Supplementary Figure 1B**).

Glycemic and blood pressure control were comparable at baseline (**Table 2**). After three diet cycles antihyperglycemic medication could be reduced in 57% of participants in the FMD group compared to 32% of the M-Diet group. In 5% of the participants of the M-Diet group antihyperglycemic medication had to be increased (**Supplementary Figure 2A**). After six diet cycles antihyperglycemic medication could be reduced in 67% of participants in the FMD group compared to baseline, whereas in 21% of the participants of the M-Diet group antihyperglycemic medication had to be increased compared to baseline (**Supplementary Figure 2A**). Antihypertensive medication could also be reduced in 10% of the participants of the FMD group compared to 5% of the participants of the M-Diet group after three and after six diet cycles. In contrast, in 5 % of the participants of the M-Diet group after three diet cycles and 10% after six diet cycles, antihypertensive medication had to be increased (**Supplementary Figure 2B**). Subjective health status, as well as somatic and depression symptoms, did not differ between the groups and throughout the study (data not shown).

Blood ketones increased 3-fold in the FMD group after three diet cycles (0.50 mmol [0.32, 0.68], P≤0.001) and after six diet cycles (0.50 mmol [0.25, 0.75], P≤0.001) (**Table 2** and **Figure 2B**). A high MDS in the M-Diet group was reported after three (6.1 MDS [5.5, 6.7]) and after six diet cycles (6.1 MDS [5.4, 6.7]) as sign of good compliance.

## Discussion

This study shows that FMD, when accompanied by intensive diabetes care, is safe and well tolerated in patients with type 2 diabetes. We found that FMD cycles can reduce ACR in patients with microalbuminuria, improve glycemic and blood pressure control and lead to a sustained improvement of HOMA-IR, even when adjusted for weight loss. Surprisingly, we observed a distinct response to FMD on markers of fatty acid oxidation in plasma of participants that responded with an improvement in ACR compared to those that did not and when compared to the M-Diet group. Exploratory analyses revealed only a transient reduction of the dicarbonyl-derived MG-H1, and of the senescence marker suPAR, whereas no difference in dicarbonyl detoxification and markers of DNA-damage/repair were observed. Interestingly, FMD-induced changes were mostly reversed after refeeding and at follow-up.

Albuminuria is thought to be the result of pathogenic events targeting the vasculature, the glomerulus, and tubular cells [35-37]. In this study we show that FMD does not change ACR levels in the overall group, but only in patients with microalbuminuria. The reduction in microalbuminuria is comparable to the reduction reported in placebo-controlled studies on SGLT-2 inhibitors [38, 39]. In contrast to the effects of SGLT-2 inhibitors, we see no difference of ACR in patients with macroalbuminuria receiving FMD, suggesting that FMD alone cannot protect once fibrosis and loss of renal architecture is established leading to a presumed point of no return. Comparable glycemic and blood pressure control between both groups, was achieved by a strong reduction in antihyperglycemic and antihypertensive medication over time in the FMD group. This reduction is attributed to the diet intervention, since no other change took place, and reflects an improved metabolic control in these patients. However, it is known that improved glycemia and blood pressure, as well as weight loss, cannot completely explain the antialbuminuric effect and other independent pathways could be involved [34].

The acylcarnitine profile and two amino acids discriminated between responders and non-responders with respect to improvement in ACR. While the specific increase in acetylcarnitine levels results most likely from pronounced activation of lipolysis during prolonged fasting periods, it is yet unclear whether acetylcarnitine is just a marker of high compliance to FMD or serves as a substrate for renal metabolism and thereby affects the response of microalbuminuria to FMD [40]. This finding is also in line with a previous report associating circulating short-chain fatty acids with normoalbuminuria in diabetes [41]. The nephroprotective effects of fatty acid oxidation and ketogenesis are probably explained by the fact that kidney metabolism relies highly on lipolysis and ketone bodies. Several studies on SGLT-2 inhibitors in diabetic patients and animal models of diabetes have shown that it is the induction of a mild ketogenesis that orchestrates the nephroprotective effects of SGLT-2 inhibitors, by shifting energy metabolism to ketolysis [23, 24]. The rapid decrease in albuminuria during FMD followed by rapid increase at refeeding points towards dynamic changes in the cellular events controlling albumin excretion. Of note, this study shows that FMD has no sustained effect on albuminuria but cannot forecast any effects on parameters determining renal function over longer time.

The increase in glycine concentration observed in the responders is in line with protective effects of glycine on renal tubular injury as previously described in experimental models of kidney injury [42]. Although we could not show a change in Glo-1 activity in WBC, previous studies have reported that glycine might enhance the function of Glo-1 and restore antioxidant defense in kidney of STZ-induced diabetic rats, thus protecting against renal oxidative stress [43].

The lack of change in plasma methylglyoxal, and in improvement of dicarbonyl detoxification was evidenced by the lack of increase in Glo-1 activity or pGlo-1 expression in WBC. Furthermore, no change in hydroxyacetone, reflecting alternative MG detoxification pathways, such as aldo-keto reductase [44], was observed in the RBC. We interpret the lack of reconstitution of DNA-damage/repair during FMD as a potential explanation for the rapid increase of ACR after termination of FMD. This is supported also by the fast increase of suPAR (Table 2) after FMD, a marker of accelerated aging and senescence [45]. Thus, none of two important defense pathways, able to control nephropathy was activated or reconstituted in this study and longer period of FMD might be needed for nephropathy reversal. However, the different response in MG-H1 reduction after three but not after six FMD cycles suggests that the changes seen are unlikely only due to reduced energy intake during FMD, and that instead a sustained redirection of metabolism is taking place during FMD to some degree.

### Strengths and limitations

Our study explores for the first time in a randomized controlled design the clinical importance of fasting in type 2 diabetes patients. No episodes of severe hypoglycemia or hypotension were reported and the intensive medical control the study participants received in this study makes on one hand this study exceptional, and on the other hand points out the complexity of a fasting intervention in type 2 diabetes patients; a message we believe will be very helpful in implementing such dietary recommendations in patient care. Moreover, our study adds to the clinical aspects, a thorough exploratory analysis. However, several limitations should be noted. First, the control group received an isocaloric diet in contrast to the caloric restriction of the FMD. Although, the control group showed a mild weight loss, weight loss in the FMD group was higher and significant when compared to baseline and to the control group. Being aware of this limitation, we adjusted all statistical analyses of the study endpoints with a correction for weight loss. Moreover, we believe that reduction of albuminuria is less likely affected by the weight loss reported in this study since such weight loss is still persistent at refeeding (one week after FMD), whereas albuminuria levels increase. Few short-term controlled studies on IF have revealed positive effects on glucose metabolism independent of weight loss [46, 47]. On the other hand, glycemia and blood pressure, two important factors that could affect albuminuria, were strictly controlled in this study, as reflected in the comparable values between the study groups throughout the study. Furthermore, we don’t see a significant change in albuminuria in the overall group after FMD treatment, but only in the subgroup analysis. The significant reduction of ACR in the subgroup analysis of patients with microalbuminuria is important, since baseline albuminuria levels have been shown to be predictors of the development of diabetic nephropathy in type 2 diabetes [48]. On the other hand, small sample size might play a role in the results of the current study and future studies should address the effect of fasting regimes in albuminuria of type 2 diabetes patients in larger cohorts, also addressing generalizability of such dietary interventions in the type 2 diabetes population. Moreover, despite a robust exploratory analysis on mechanistic explanations of the reduction in ACR levels, our study is limited in exploring renal cell metabolism as we did not collect kidney biopsies in this study.

## Conclusion

This proof-of-concept-study showed that when accompanied by intensive diabetes care, FMD cycles can be well integrated into clinical practice complementary to current guidelines and that it can lead to a reduction in ACR in type 2 diabetes patients with microalbuminuria, an improvement of glycemic and blood pressure control and to a sustained reduction in insulin resistance also when adjusted for weight loss. Lack of changes in markers of dicarbonyl detoxification and of DNA-damage/repair might explain the relapse of albuminuria after six FMD cycles, suggesting that FMD cycles may have to be continued for longer periods. Distinct effects of FMD on markers of incomplete lipid oxidation in patients that show an improvement of albuminuria, might help in the future to develop new therapeutic strategies.

## Data Availability

The data generated in this study are subject to national data protection laws and are available from the corresponding author upon reasonable request.

## Abbreviations

AC: acylcarnitines
ACR: albumin-to-creatinine ratio
C2: acetylcarnitine
CTCAE: common terminology criteria for adverse events
DKD: diabetic kidney disease
FMD: fasting-mimicking diet
FPG: fasting plasma glucose
Glo-1: glyoxalase-1
MDS: Mediterranean diet score
MG: methylglyoxal
MG-H1: methylglyoxal-derived hydroimidazolone 1
RBC: red blood cells
PHQ: patient health questionnaire
SF-12: 12-item short-form health survey questionnaire
SGLT2: sodium-glucose cotransporter-2
pGlo-1: phosphorylated glyoxalase-1
WBC: white blood cells

## Acknowledgments

The authors thank all study participants and staff members of the Study Ambulance for Diabetes Research Heidelberg involved in the conduct of the study. We thank you Marietta Kirchner (Institute of Medical Biometry, Heidelberg), for her assistance in the statistical analysis.

## Author Contribution

A.S., E.R., E.K., Z.K., H.B., M.B., F.G.C., Z.H., V.K., J.G.O., J.M., and T.F. contributed to the acquisition and interpretation of data. A.S., S.H. and P.N. contributed to the study design and interpretation of data. A.S., J.S. and P.N. drafted the report. All authors contributed to the review of the report and approved the final version for submission. A.S. and P.N. are the guarantors of this work and, as such, had full access to all the data in the study and takes responsibility for the integrity of the data and the accuracy of the data analysis.

