## Supplementary material for "A six-month periodic fasting reduces microalbuminuria and improves metabolic control in patients with type 2 diabetes and diabetic nephropathy: a randomized controlled study"

**SUPPLEMENTARY DATA**

**Supplementary Table S1. Diet composition.**

**(A)** Fasting-mimicking diet composition.

| | Day 1 | Day 2-5 |
|---|---|---|
| Energy (kJ/day)<br>Energy (Cal/day) | 4600<br>1099 | 3000<br>717 |
| Protein (%)<br>Fat (%)<br>Carbohydrates (%) | 11<br>46<br>43 | 9<br>44<br>47 |

**(B)** Dietary recommendation to patients in the Mediterranean-diet group.

| Mediterranean Diet Score | | |
|---|---|---|
| Food recommended | Recommendation | Yes=1 / No=0 |
| Olive oil (alternative Canola oil) | 2 tbsp/day | |
| Tree nuts | max. 1 handfull (25g)/day | |
| Bread<br>potatoe, pasta, rice | 1 slice (70-85g)/day,<br>1-2 handfull (200-250)/day<br>(whole grain preferred) | |
| Fresh fruits | 2 servings (125g)/day<br>1-2 handfull/day | |
| Vegetables, green salads | at least 2 servings (140g)/day<br>at least 1-2 handfull/day | |
| Fish, seafood | 2-3 servings (150-200g)/week | |
| Legumes | 2-3 servings (200g)/week | |
| White meat | 1-2 servings (150-200g)/week | |
| Wine | Moderate consume with meals (100ml/day), optionally only for habitual drinkers | |
| MDS total | | 0-9 |

| Food discouraged | Recommendation |
|---|---|
| Soda drinks | No consume |
| Commercial bakery goods, sweets, pastries | Max. 3 servings (1 piece, 1 handful, 220-270 Cal)/week |
| Red meat<br>Sausages | Max. 1 serving (150-200 g)/week<br>Max. 1 serving (30 g)/week |
| Spread fats (butter/margarine) | Max. 1 serving (10 g)/day |

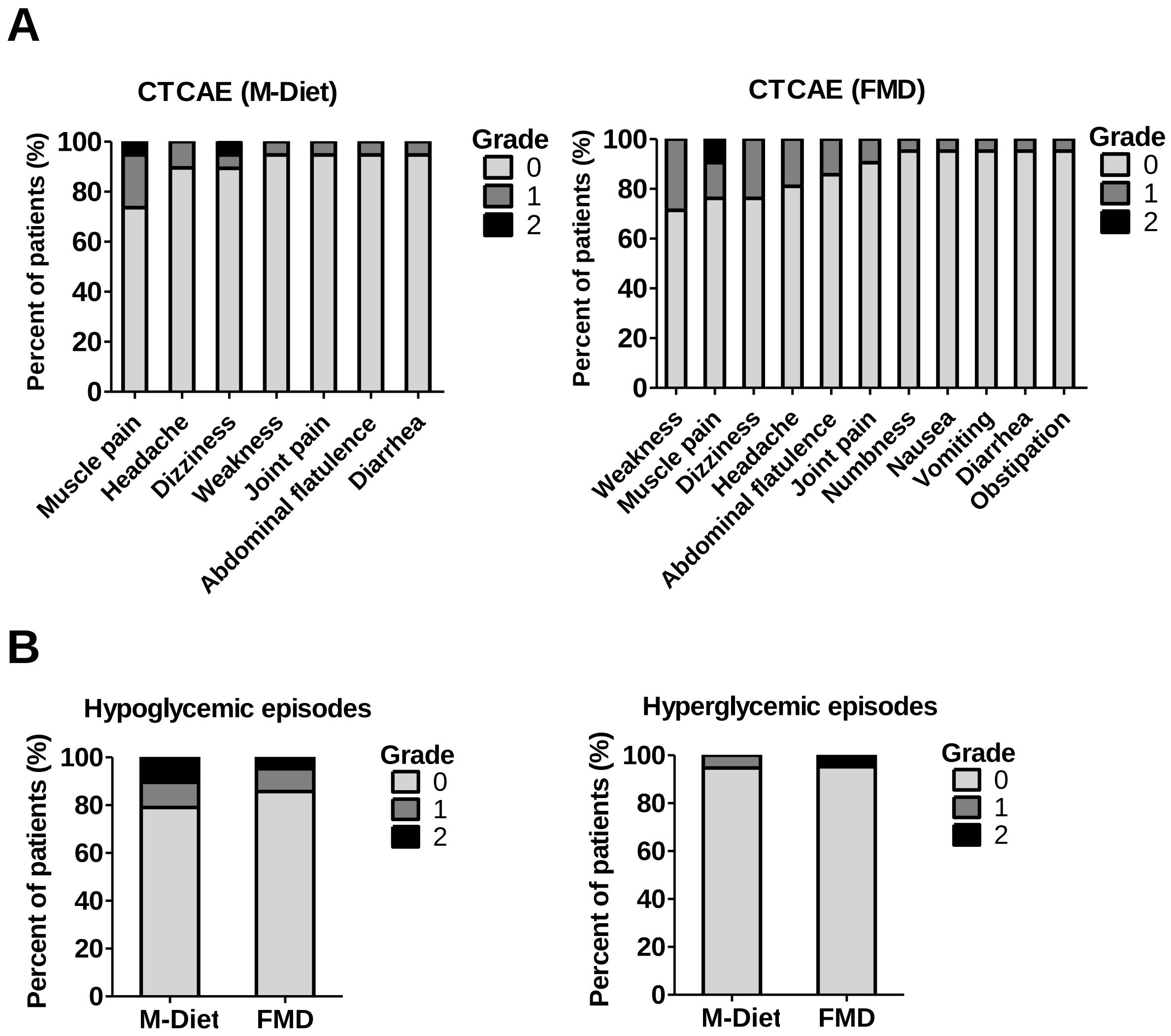


**Supplementary Figure 1.** Subject self-reported adverse effects based on CTCAE (A). Hypoglycemic and hyperglycemic episodes in both study groups reported during the study (B). (CTCAE; 1 = Mild, 2 = Moderate, 3 = Severe, 4 = Life-threatening, 5 = Death). Percentage of subjects reporting no adverse effect (grade 0), grade 1 or grade 2 adverse effects; grades 3 to 5 were not reported. Adverse effects were assessed following Common Terminology Criteria for Adverse Events (CTCAE).
M-Diet Mediterranean Diet, FMD fasting-mimicking diet.

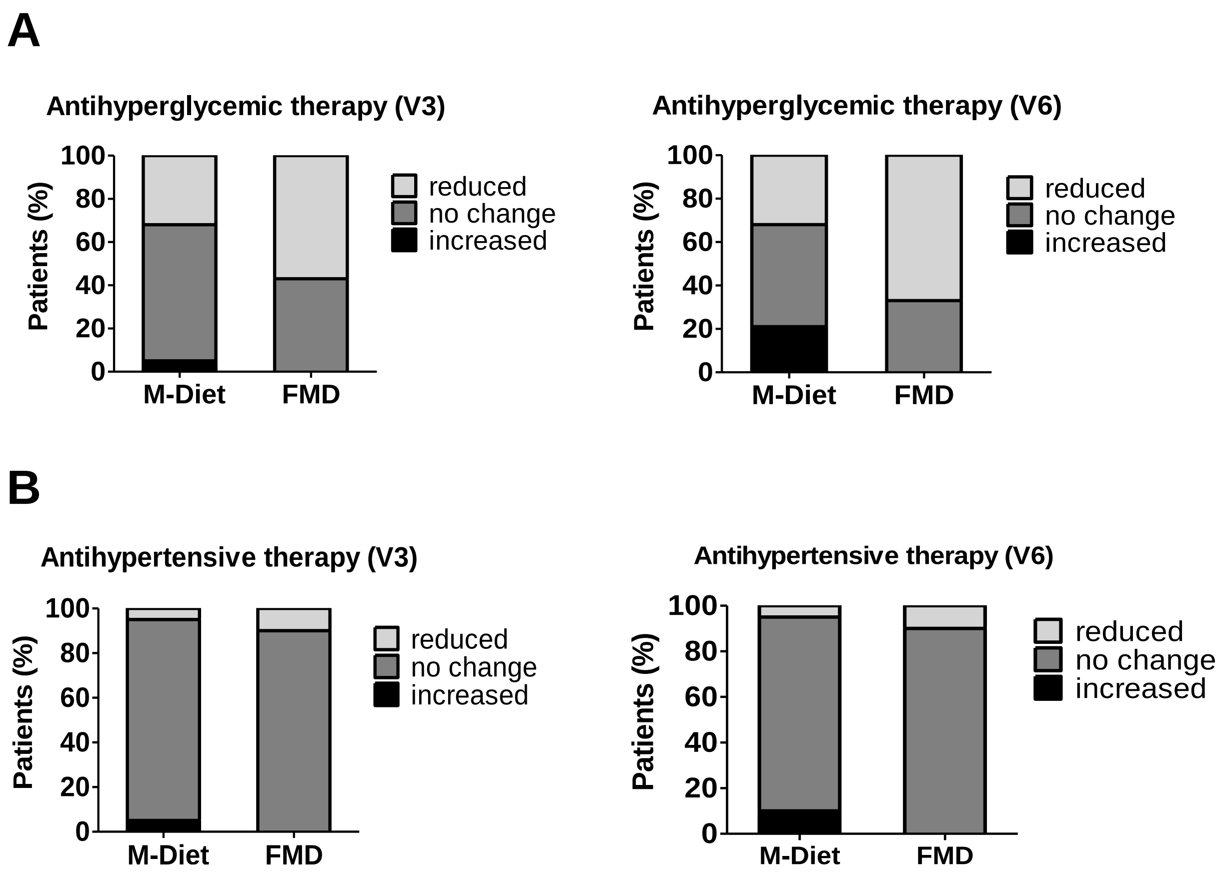


**Supplementary Figure 2.** Change of antihyperglycemic (A) and antihypertensive medication (B) after 3 diet cycles (V3) and after 6 diet cycles (V6).
M-Diet Mediterranean diet, FMD fasting-mimicking diet.

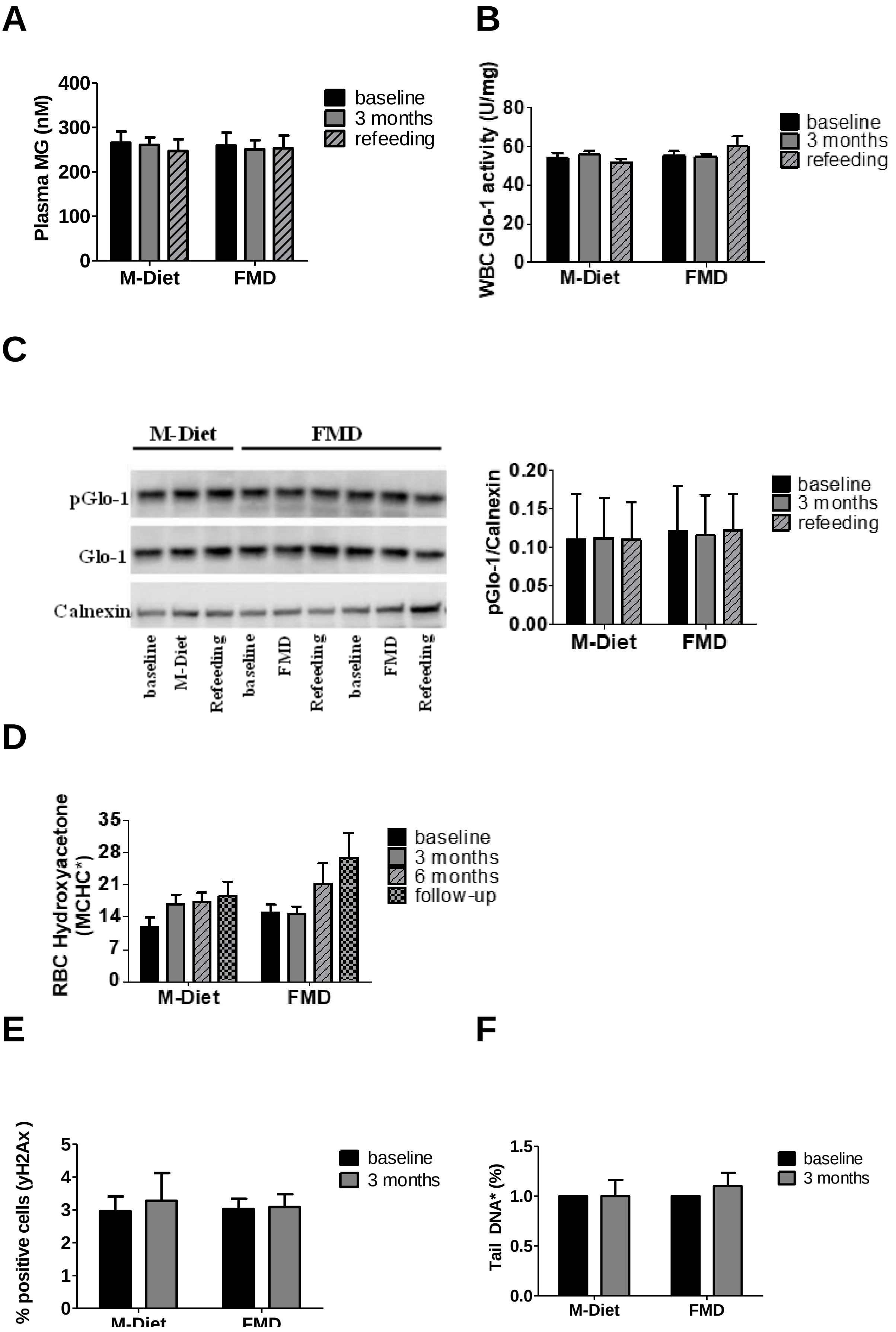
A
Plasma MG (nM)
400
300
200
100
0
M-Diet
FMD
baseline
3 months
refeeding
B
WBC Glo-1 activity (U/mg)
80
60
40
20
0
M-Diet
FMD
baseline
3 months
refeeding
C
M-Diet
FMD
pGlo-1
Glo-1
Calnexin
baseline
M-Diet
Refeeding
baseline
FMD
Refeeding
baseline
FMD
Refeeding
pGlo-1/Calnexin
0.20
0.15
0.10
0.05
0.00
M-Diet
FMD
baseline
3 months
refeeding
D
RBC Hydroxyacetone (MCHC*)
35
28
21
14
7
0
M-Diet
FMD
baseline
3 months
6 months
follow-up
E
% positive cells (yH2Ax )
5
4
3
2
1
0
M-Diet
FMD
baseline
3 months
F
Tail DNA* (%)
1.5
1.0
0.5
0.0
M-Diet
FMD
baseline
3 months

**Supplementary Figure 3.** Plasma methylglyoxal level (A), glyoxalase 1 activity in white blood cells (B), phosphorylated glyoxalase-1 expression in white blood cells (C), hydroxyacetone concentration in red blood cells (D), yH2Ax expression in white blood cells (E), comet assay with white blood cells (F)

Data are shown as mean ± SEM of unadjusted values of parameter. Total Glo-1 immunoblotting is a rehybridization of the pGlo-1 in (C).

Representative participants were analyzed in (A) M-Diet n=11, FMD n=11; (B) M-Diet n=11, FMD n=12; (C) M-Diet n=6, FMD n=12; (D) M-Diet n=19, FMD n=21; (E) M-Diet n=6, FMD n=9; (F) M-Diet n=6, FMD n=12.

Abbreviations: M-Diet Mediterranean diet, FMD fasting-mimicking diet, MG methylglyoxal, WBC white blood cell, Glo-1 glyoxalase 1, pGlo-1 phosphorilated glyoxalase 1, RBC red blood cells, MCHC mean corpuscular hemoglobin concentration, yH2Ax phosphorylated histone H2AX.

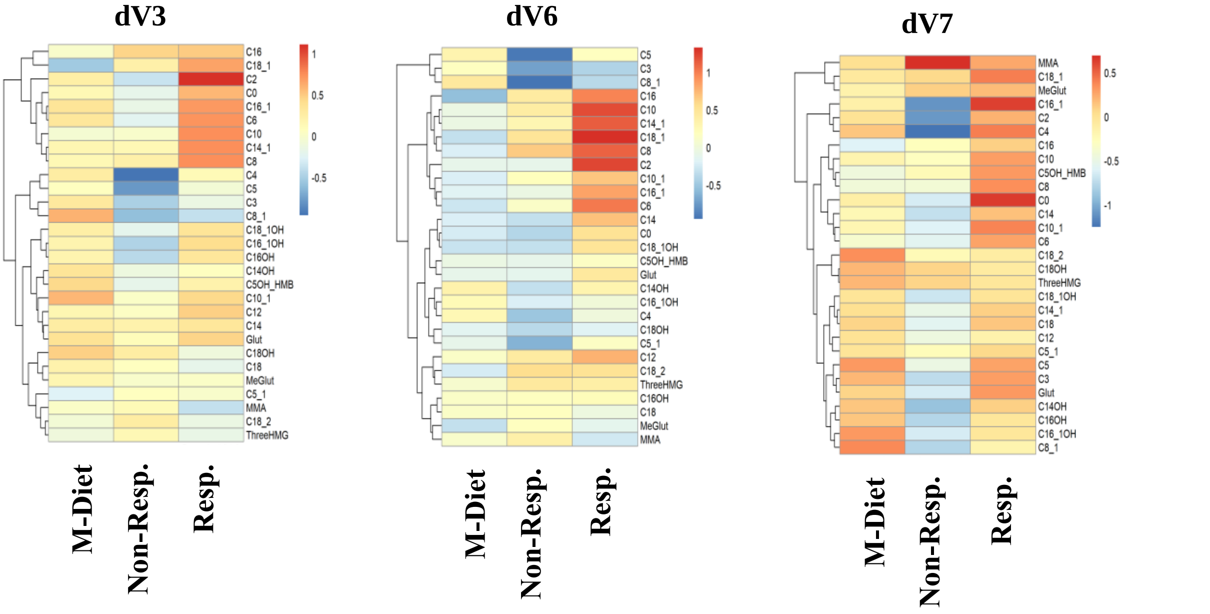


**Supplementary Figure 4.** Effects of fasting on acylcarnitines after 3 months, after 6 months and at follow-up.

Heat map analysis of acylcarnitine profile reveal differences between the study groups dependent on change in albuminuria. Patient in the intervention group that show at least a 30% decrease in albuminuria level after 3 and after 6 diet cycles compared to the respective baseline are referred as Responders, while the rest are referred as non-Responders. Groups in the analysis: M-Diet, Non-Responders and Responders. Each row displays a metabolite and each column represent the absolute change of the annotated metabolite after 3 diet cycles (left panel), after 6 diet cycles (middle panel) and at follow-up (right panel) compared to baseline of the respective group and is displayed as range-scaled Z-score. Metabolites increased are displayed in red while metabolites decreased are displayed in blue.

Abbreviations: M-Diet Mediterranean diet, Non-Resp. non-responders, Resp. responders, C0 Carnitine, C2 Acetylcarnitine, C3 Propionylcarnitine, C4 Butyrylcarnitine, Isobutyrylcarnitine, C5_1 Tiglylcarnitine, Methylcrotonylcarnitine, C5 Valerylcarnitine, Isovalerylcarnitine, Methylbutyrylcarnitine, C6 Hexanoylcarnitine, C5OH+HMB Hydroxyvalerylcarnitine+2-OH-3-Methyl-butyrylcarnitin, C8_1 Octenoylcarnitine, C8Octanoylcarnitine, C10_1 Decenoylcarnitine, C10 Decanoylcarnitine, MMA Methylmalonylcarnitin, Glut Glutarylcarnitine, C12 Dodecanoylcarnitine, MeGlut Methylglutarylcarnitine, 3HMG 3-Hydroxy-3-methylglutarylcarnitin, C14_1 Tetradecenoylcarnitine, C14 Tetradecanoylcarnitine, C14OH Hydroxytetradecanoylcarnitin, C16_1 Hexadecenoylcarnitine, C16 Hexadecanoylcarnitine, C16_1OH Hydroxyhexadecenoylcarnitine, C16OH Hydroxyhexadecanoylcarnitine, C18_2 Octadecadienylcarnitine, C18_1 Octadecenoylcarnitine, C18 Octadecanoylcarnitine, C18_1OH Hydroxyoctadecenoylcarnitine, C18OH Hydroxyoctadecanoylcarnitin.

**Supplementary Table S2.** Acylcarnitine profile and amino acids at baseline, after 3 diet cycles (V3), after 6 diet cycles (V6) and at follow-up (V7)

| **Parameter Group** | | **V0** | **V3** | **V6** | **V7** |
|---|---|---|---|---|---|
| C0 | M-Diet | 32.64±1.75 (18) | 33.23±1.91 (18) | 30.53±1.74 (16) | 30.31±1.39 (15) |
| | Non-Resp. | 34.74±4.62 (9) | 33.02±4.14 (9) | 31.29±4.28 (7) | 31.79±3.25 (7) |
| | Resp. | 30.92±2.33 (11) | 34.54±2.58 (11) | 32.69±3.90 (10) | 34.84±3.78 (9) |
| **C2** | M-Diet | 8.19±0.60 (18) | 8.87±1.11 (18) | 7.53±0.62 (16) | 7.37±0.67 (15) |
| | Non-Resp. | 11.98±1.74 (9) | 10.12±0.80 (9) | 11.22±0.78 (7) | 7.87±1.34 (7) |
| | Resp. | 8.94±1.06 (11) | **13.11±1.97 (11)*** | **12.82±1.25 (10)**** | 9.12±1.07 (9) |
| C3 | M-Diet | 0.41±0.04 (18) | 0.44±0.04 (18) | 0.42±0.04 (16) | 0.4±0.03 (15) |
| | Non-Resp. | 0.54±0.08 (9) | 0.43±0.07 (9) | 0.38±0.06 (7) | 0.44±0.08 (7) |
| | Resp. | 0.37±0.04 (11) | 0.32±0.04 (11) | 0.28±0.04 (10) | 0.43±0.07 (9) |
| C4 | M-Diet | 0.21±0.02 (18) | 0.22±0.03 (18) | 0.21±0.03 (16) | 0.18±0.01 (15) |
| | Non-Resp. | 0.27±0.07 (9) | 0.16±0.03 (9) | 0.14±0.02 (7) | 0.16±0.02 (7) |
| | Resp. | 0.2±0.03 (11) | 0.19±0.02 (11) | 0.19±0.03 (10) | 0.23±0.02 (9) |
| C5:1 | M-Diet | 0.02±0.00 (18) | 0.01±0.00 (18) | 0.02±0.00 (16) | 0.02±0.00 (15) |
| | Non-Resp. | 0.03±0.00 (9) | 0.02±0.01 (9) | 0.01±0.00 (7) | 0.02±0.00 (7) |
| | Resp. | 0.02±0.00 (11) | 0.02±0.00 (11) | 0.02±0.00 (10) | 0.02±0.00 (9) |
| **C5** | M-Diet | 0.14±0.01 (18) | 0.12±0.01 (18) | 0.13±0.01 (16) | 0.12±0.01 (15) |
| | Non-Resp. | 0.17±0.02 (9) | 0.11±0.02 (9) | 0.08±0.01 (7) | 0.14±0.03 (7) |
| | Resp. | 0.13±0.01 (11) | 0.11±0.01 (11) | **0.12±0.01 (10)*** | 0.13±0.01 (9) |
| **C6** | M-Diet | 0.07±0.01 (18) | 0.1±0.02 (18) | 0.07±0.01 (16) | 0.07±0.01 (15) |
| | Non-Resp. | 0.1±0.01 (9) | 0.1±0.01 (9) | 0.11±0.02 (7) | 0.08±0.01 (7) |
| | Resp. | 0.07±0.01 (11) | 0.11±0.02 (11) | **0.12±0.02 (10)*** | 0.09±0.02 (9) |
| C5OH + HMB | M-Diet | 0.04±0 (16) | 0.05±0.01 (16) | 0.04±0 (9) | 0.03±0.01 (9) |
| | Non-Resp. | 0.05±0 (9) | 0.04±0.01 (9) | 0.04±0.01 (5) | 0.04±0.01 (5) |
| | Resp. | 0.04±0.01 (8) | 0.04±0 (8) | 0.04±0.01 (6) | 0.04±0.01 (6) |
| **C8:1** | M-Diet | 0.23±0.03 (18) | 0.26±0.03 (18) | 0.24±0.05 (16) | 0.21±0.04 (15) |
| | Non-Resp. | 0.41±0.09 (9) | 0.25±0.04 (9) | 0.18±0.03 (7) | 0.3±0.09 (7) |
| | Resp. | 0.31±0.04 (11) | **0.17±0.02 (11)**** | **0.18±0.02 (10)*** | 0.22±0.04 (9) |
| **C8** | M-Diet | 0.13±0.02 (18) | 0.17±0.03 (18) | 0.14±0.03 (16) | 0.13±0.02 (15) |
| | Non-Resp. | 0.14±0.02 (9) | 0.18±0.02 (9) | 0.22±0.02 (7) | 0.13±0.02 (7) |
| | Resp. | 0.14±0.02 (11) | 0.23±0.05 (11) | **0.25±0.05 (10)**** | 0.19±0.04 (9) |
| C10:1 | M-Diet | 0.23±0.03 (18) | 0.36±0.07 (18) | 0.23±0.04 (16) | 0.23±0.04 (15) |
| | Non-Resp. | 0.3±0.03 (9) | 0.33±0.04 (9) | 0.35±0.05 (7) | 0.23±0.04 (7) |
| | Resp. | 0.23±0.04 (11) | 0.33±0.07 (11) | 0.36±0.07 (10) | 0.33±0.07 (9) |
| **C10** | M-Diet | 0.17±0.02 (18) | 0.23±0.04 (18) | 0.23±0.03 (16) | 0.21±0.02 (15) |
| | Non-Resp. | 0.21±0.04 (9) | 0.27±0.03 (9) | 0.36±0.04 (7) | 0.22±0.03 (7) |
| | Resp. | 0.23±0.03 (11) | 0.40±0.11 (11) | **0.47±0.09 (10)*** | 0.32±0.08 (9) |
| MMA | M-Diet | 0.06±0.01 (16) | 0.06±0.01 (16) | 0.07±0.01 (9) | 0.07±0.01 (9) |
| | Non-Resp. | 0.06±0.01 (9) | 0.07±0 (9) | 0.06±0.01 (5) | 0.07±0.01 (5) |
| | Resp. | 0.06±0.01 (8) | 0.05±0.01 (8) | 0.05±0.01 (6) | 0.07±0.02 (6) |

| | | | | | |
|---|---|---|---|---|---|
| Glut | M-Diet | 0.2±0.01 (18) | 0.22±0.02 (18) | 0.18±0.02 (16) | 0.18±0.02 (15) |
| | Non-Resp. | 0.22±0.03 (9) | 0.22±0.02 (9) | 0.2±0.03 (7) | 0.18±0.02 (7) |
| | Resp. | 0.2±0.03 (11) | 0.23±0.04 (11) | 0.23±0.04 (10) | 0.23±0.04 (9) |
| C12 | M-Diet | 0.08±0.01 (18) | 0.09±0.01 (18) | 0.09±0.01 (16) | 0.08±0.01 (15) |
| | Non-Resp. | 0.11±0.02 (9) | 0.11±0.01 (9) | 0.14±0.02 (7) | 0.09±0.02 (7) |
| | Resp. | 0.11±0.01 (11) | 0.14±0.03 (11) | 0.15±0.03 (10) | 0.11±0.02 (9) |
| MeGlut | M-Diet | 0.06±0.01 (18) | 0.05±0.01 (18) | 0.04±0.01 (16) | 0.04±0.01 (15) |
| | Non-Resp. | 0.05±0.01 (9) | 0.04±0.01 (9) | 0.04±0.01 (7) | 0.05±0.01 (7) |
| | Resp. | 0.05±0.01 (11) | 0.05±0.01 (11) | 0.04±0.01 (10) | 0.06±0.02 (9) |
| 3HMG | M-Diet | 0.01±0.00 (18) | 0.01±0.00 (18) | 0.01±0.00 (16) | 0.01±0.00 (15) |
| | Non-Resp. | 0.01±0.00 (9) | 0.01±0 (9) | 0.01±0.01 (7) | 0.01±0.00 (7) |
| | Resp. | 0.01±0.00 (11) | 0.01±0.00 (11) | 0.02±0.00 (10) | 0.01±0.00 (9) |
| **C14:1** | M-Diet | 0.07±0.01 (18) | 0.1±0.01 (18) | 0.08±0.01 (16) | 0.08±0.01 (15) |
| | Non-Resp. | 0.11±0.02 (9) | 0.13±0.02 (9) | 0.16±0.03 (7) | 0.09±0.01 (7) |
| | Resp. | 0.1±0.02 (11) | 0.17±0.05 (11) | **0.19±0.05 (10)*** | 0.12±0.02 (9) |
| C14 | M-Diet | 0.04±0.00 (18) | 0.05±0.00 (18) | 0.03±0.00 (16) | 0.03±0.00 (15) |
| | Non-Resp. | 0.05±0.01 (9) | 0.05±0.01 (9) | 0.04±0.00 (7) | 0.03±0.01 (7) |
| | Resp. | 0.04±0.01 (11) | 0.05±0.01 (11) | **0.06±0.01 (10)*** | 0.05±0.01 (9) |
| C14OH | M-Diet | 0.03±0.00 (18) | 0.04±0.01 (18) | 0.04±0.01 (16) | 0.03±0.01 (15) |
| | Non-Resp. | 0.06±0.01 (9) | 0.05±0.00 (9) | 0.05±0.01 (7) | 0.02±0.00 (7) |
| | Resp. | 0.04±0.01 (11) | 0.03±0.01 (11) | 0.04±0.01 (10) | 0.04±0.01 (9) |
| **C16:1** | M-Diet | 0.02±0.00 (18) | 0.03±0.00 (18) | 0.02±0.00 (16) | 0.02±0.00 (15) |
| | Non-Resp. | 0.03±0.00 (9) | 0.03±0.00 (9) | 0.03±0.01 (7) | 0.02±0.00 (7) |
| | Resp. | 0.02±0.00 (11) | 0.03±0.01 (11) | **0.03±0.00 (10)*** | 0.03±0.01 (9) |
| **C16** | M-Diet | 0.12±0.01 (18) | 0.13±0.01 (18) | 0.10±0.01 (16) | 0.1±0.01 (15) |
| | Non-Resp. | 0.12±0.01 (9) | 0.15±0.02 (9) | 0.14±0.01 (7) | 0.11±0.01 (7) |
| | Resp. | 0.12±0.02 (11) | 0.14±0.02 (11) | **0.16±0.02 (10)**** | 0.12±0.02 (9) |
| C16:1 OH | M-Diet | 0.01±0.00 (18) | 0.01±0.00 (18) | 0.01±0.00 (16) | 0.01±0.00 (15) |
| | Non-Resp. | 0.02±0.00 (9) | 0.01±0.00 (9) | 0.01±0.00 (7) | 0.01±0.00 (7) |
| | Resp. | 0.01±0.00 (11) | 0.02±0.00 (11) | 0.01±0.00 (10) | 0.01±0.00 (9) |
| C16OH | M-Diet | 0.01±0.00 (18) | 0.01±0.00 (18) | 0.01±0.00 (16) | 0.01±0.00 (15) |
| | Non-Resp. | 0.02±0.00 (9) | 0.02±0.00 (9) | 0.02±0.01 (7) | 0.01±0.00 (7) |
| | Resp. | 0.01±0.00 (11) | 0.02±0.00 (11) | 0.01±0.00 (10) | 0.01±0.00 (9) |
| C18:2 | M-Diet | 0.03±0.00 (18) | 0.03±0.00 (18) | 0.03±0.00 (16) | 0.04±0.00 (15) |
| | Non-Resp. | 0.04±0.01 (9) | 0.04±0.00 (9) | 0.05±0.01 (7) | 0.03±0.00 (7) |
| | Resp. | 0.04±0.01 (11) | 0.04±0.01 (11) | 0.05±0.01 (10) | 0.04±0.01 (9) |
| **C18:1** | M-Diet | 0.07±0.00 (18) | 0.06±0.00 (18) | 0.07±0.01 (16) | 0.08±0.01 (15) |
| | Non-Resp. | 0.07±0.01 (9) | 0.09±0.01 (9) | 0.10±0.01 (7) | 0.08±0.01 (7) |
| | Resp. | 0.08±0.01 (11) | **0.11±0.02 (11)*** | **0.13±0.02 (10)**** | 0.1±0.01 (9) |
| C18 | M-Diet | 0.05±0.00 (18) | 0.06±0.00 (18) | 0.05±0.01 (16) | 0.05±0.01 (15) |
| | Non-Resp. | 0.06±0.01 (9) | 0.06±0.00 (9) | 0.06±0.01 (7) | 0.04±0.01 (7) |
| | Resp. | 0.06±0.01 (11) | 0.06±0.01 (11) | 0.06±0.01 (10) | 0.07±0.01 (9) |
| | M-Diet | 0.01±0.00 (18) | 0.01±0.00 (18) | 0.01±0.00 (16) | 0.01±0.00 (15) |

| | | | | | |
|---|---|---|---|---|---|
| C18:1 OH | Non-Resp. | 0.02±0.00 (9) | 0.01±0.00 (9) | 0.01±0.00 (7) | 0.01±0.00 (7) |
| | Resp. | 0.01±0.00 (11) | 0.01±0.00 (11) | 0.01±0.00 (10) | 0.01±0.00 (9) |
| C18OH | M-Diet | 0.01±0.00 (18) | 0.01±0.00 (18) | 0.01±0.00 (16) | 0.01±0.00 (15) |
| | Non-Resp. | 0.01±0.00 (9) | 0.01±0.00 (9) | 0.01±0.00 (7) | 0.02±0.00 (7) |
| | Resp. | 0.01±0.00 (11) | 0.01±0.00 (11) | 0.01±0.00 (10) | 0.01±0.00 (9) |
| **Ala** | M-Diet | 238.2±11.65 (18) | 238.19±13.09 (18) | 238.86±9.48 (16) | 238.8±13.49 (15) |
| | Non-Resp. | 226.76±17.24 (9) | 218.95±26.1 (9) | 238.26±28.68 (7) | 291.39±25.37 (7) |
| | Resp. | 232.23±14.31 (11) | 201.8±15.88 (11) | **189.14±13.56 (10)*** | 226.3±20.89 (9) |
| Pro | M-Diet | 737.72±95.58 (18) | 687.26±69.3 (18) | 488.52±74.99 (16) | 548.9±95.78 (15) |
| | Non-Resp. | 817.94±103.61 (9) | 671.79±78.23 (9) | 464.34±88.37 (7) | 828.44±193.11 (7) |
| | Resp. | 573.94±90.03 (11) | 449.57±79.73 (11) | 383.68±73.77 (10) | 499.78±93.77 (9) |
| **Val** | M-Diet | 184.28±4.72 (18) | 184.2±5.04 (18) | 197.86±7.75 (16) | 195.58±9.98 (15) |
| | Non-Resp. | 203.3±13.45 (9) | 164.02±4.77 (9) | 181.78±7.33 (7) | 222.8±14.64 (7) |
| | Resp. | 183.77±7.95 (11) | **157.73±6.69 (11)*** | 173.79±6.46 (10) | 181.11±9.4 (9) |
| Thr | M-Diet | 99.19±12.11 (18) | 92.93±9.87 (18) | 60.86±12.36 (16) | 66.13±13.9 (15) |
| | Non-Resp. | 90.95±7.18 (9) | 102.55±9.79 (9) | 77.14±20.48 (7) | 90.4±24.31 (7) |
| | Resp. | 71.72±13.77 (11) | 83.7±15.75 (11) | 74.31±18.78 (10) | 78.47±18.14 (9) |
| Leu/Ile | M-Diet | 138.34±4.78 (18) | 142.77±5.76 (18) | 171.19±12.79 (16) | 168.15±12.7 (15) |
| | Non-Resp. | 153.77±8.67 (9) | 140.93±8.86 (9) | 160.06±9.26 (7) | 182.62±11.37 (7) |
| | Resp. | 149.91±9.78 (11) | 135.03±11.91 (11) | 158.76±13.28 (10) | 145.99±12.07 (9) |
| Gln | M-Diet | 614.0±228.1 (18) | 652.4±238.9 (18) | 1597.8±390.5 (16) | 1386.3±370.1 (15) |
| | Non-Resp. | 282.9±20.7 (9) | 285.1±19.1 (9) | 1170.6±577.5 (7) | 1197.58±588.68 (7) |
| | Resp. | 1119.9±442.8 (11) | 1132.4±434.0 (11) | 1499.4±488.9 (10) | 1247.63±475.48 (9) |
| Met | M-Diet | 18.81±1.11 (18) | 19.23±1.12 (18) | 18.64±1.13 (16) | 19.53±1.01 (15) |
| | Non-Resp. | 16.83±1.8 (9) | 19.52±1.45 (9) | 22.14±1.44 (7) | 19.28±2.1 (7) |
| | Resp. | 17.93±1 (11) | 20.45±1.47 (11) | 20.23±1.17 (10) | 19.14±1.21 (9) |
| His | M-Diet | 1093.3±76 (18) | 1206.6±98.6 (18) | 1381.1±109.9 (16) | 1248.5±117.1 (15) |
| | Non-Resp. | 979.9±135.11 (9) | 1063.7±148.4 (9) | 1187.1±137.6 (7) | 1279.9±139.0 (7) |
| | Resp. | 1201.1±120.3 (11) | 1263.7±180.7 (11) | 1161.2±117.3 (10) | 1103.1±102.6 (9) |
| Phe | M-Diet | 42.58±1.15 (18) | 43.34±1.39 (18) | 48.72±1.82 (16) | 48.3±2.42 (15) |
| | Non-Resp. | 40.67±1.61 (9) | 40.43±1.31 (9) | 46.86±3.83 (7) | 47.56±2.15 (7) |
| | Resp. | 43.84±2.1 (11) | 45.32±2.66 (11) | 46.73±2.58 (10) | 45.75±2.31 (9) |
| Tyr | M-Diet | 61.9±3.33 (18) | 62.07±3.05 (18) | 64.94±3.5 (16) | 62.46±3.19 (15) |

| | | | | | |
|---|---|---|---|---|---|
| | Non-Resp. | 60.39±6.77 (9) | 57.74±2.7 (9) | 58.65±5.4 (7) | 68.55±8.6 (7) |
| | Resp. | 64.31±5.62 (11) | 58.77±4.44 (11) | 57.09±3.14 (10) | 59.84±3.29 (9) |
| Asp | M-Diet | 45.71±2.73 (18) | 44.39±2.21 (18) | 38.09±2.88 (16) | 40.42±3.57 (15) |
| | Non-Resp. | 46.46±3.12 (9) | 40.92±3.88 (9) | 40.28±3.93 (7) | 43.81±5.96 (7) |
| | Resp. | 40.3±2.46 (11) | 42.12±3.84 (11) | 37.33±2.49 (10) | 41.88±4.62 (9) |
| Glu | M-Diet | 136.56±8.34 (18) | 118.8±4.75 (18) | 104.18±9.5 (16) | 101.36±10.16 (15) |
| | Non-Resp. | 115.64±10.09 (9) | 112.81±7.76 (9) | 98.7±13.51 (7) | 97.31±12.91 (7) |
| | Resp. | 126.91±19.35 (11) | 110.13±10.29 (11) | 88.97±10.01 (10) | 109.78±16.53 (9) |
| Trp | M-Diet | 53.92±12.18 (18) | 52.75±13.21 (18) | 124.59±26.71 (16) | 128.86±32.02 (15) |
| | Non-Resp. | 32±1.61 (9) | 28.73±1.6 (9) | 103.36±51.08 (7) | 105.34±44.83 (7) |
| | Resp. | 97.44±31.64 (11) | 99.88±33.67 (11) | 110.51±34.63 (10) | 107.67±36.67 (9) |
| **Gly** | M-Diet | 152.41±5.78 (18) | 157.72±6.01 (18) | 147.37±9.64 (16) | 145.69±8.46 (15) |
| | Non-Resp. | 176.08±16.04 (9) | 197.41±14.38 (9) | 193.34±21.57 (7) | 197.12±17.59 (7) |
| | Resp. | 169.65±10.6 (11) | **213.02±16.23 (11)*** | **209.02±17.65 (10)*** | 159.82±12.36 (9) |
| Orn | M-Diet | 31.56±2.55 (18) | 30.31±2.27 (18) | 42.12±4.5 (16) | 38.61±4.48 (15) |
| | Non-Resp. | 28.51±2.27 (9) | 31.82±4.56 (9) | 32.96±5.07 (7) | 38.83±6.38 (7) |
| | Resp. | 36.34±5.94 (11) | 37.14±4.7 (11) | 43.77±7.06 (10) | 42.52±7.88 (9) |
| Arg | M-Diet | 57.68±4.86 (18) | 53.99±4.31 (18) | 63.26±5.54 (16) | 58.9±5.02 (15) |
| | Non-Resp. | 47.54±3.01 (9) | 55.14±4.99 (9) | 58.63±5.93 (7) | 65.35±8.72 (7) |
| | Resp. | 60.64±8.16 (11) | 67.62±3.51 (11) | 66.01±5.73 (10) | 64.93±4.95 (9) |
| Cit | M-Diet | 21.43±1.99 (18) | 23.46±1.67 (18) | 25.62±2.11 (16) | 23.39±1.79 (15) |
| | Non-Resp. | 21.78±2.91 (9) | 25.99±3.7 (9) | 24.02±3.85 (7) | 22.56±3.45 (7) |
| | Resp. | 23.09±3.57 (11) | 26.73±2.75 (11) | 28.19±2.56 (10) | 29.82±4.48 (9) |
| Hci | M-Diet | 1.66±0.25 (18) | 1.62±0.21 (18) | 1.41±0.26 (16) | 1.4±0.22 (15) |
| | Non-Resp. | 2.28±0.43 (9) | 1.68±0.32 (9) | 1.56±0.41 (7) | 1.63±0.36 (7) |
| | Resp. | 1.36±0.26 (11) | 1.16±0.19 (11) | 1.27±0.23 (10) | 1.25±0.24 (9) |
| Asa | M-Diet | 0.2±0.05 (18) | 0.18±0.03 (18) | 0.18±0.04 (16) | 0.18±0.07 (15) |
| | Non-Resp. | 0.15±0.01 (9) | 0.16±0.01 (9) | 0.21±0.08 (7) | 0.2±0.04 (7) |
| | Resp. | 0.14±0.01 (11) | 0.13±0.02 (11) | 0.14±0.03 (10) | 0.09±0.02 (9) |

Data are shown as mean ± SEM of unadjusted values of variable. Patient in the intervention group that show at least a 30% decrease in albuminuria compared to the respective baseline are referred as Responders, while the rest are referred as non-Responders. Statistical significant values are written in bold and indicate significance level of intervention effect on change of parameter compared to baseline and are based on M-Diet corrected ANCOVA with adjustment for age, sex and weight loss. *** P≤0.001, **P≤0.01, *P≤0.05. Unit of measurement is μmol/L.

*Abbreviations:* C0 Carnitine, C2 Acetylcarnitine, C3 Propionylcarnitine, C4 Butyrylcarnitine, Isobutyrylcarnitine, C5_1 Tiglylcarnitine, Methylcrotonylcarnitine, C5

Valerylcarnitine, Isovalerylcarnitine, Methylbutyrylcarnitine, C6 Hexanoylcarnitine, C5OH+HMB Hydroxyvalerylcarnitine+2-OH-3-Methyl-butyrylcarnitin, C8_1 Octenoylcarnitine, C8Octanoylcarnitine, C10_1 Decenoylcarnitine, C10 Decanoylcarnitine, MMA Methylmalonylcarnitin, Glut Glutarylcarnitine, C12 Dodecanoylcarnitine, MeGlut Methylglutarylcarnitine, 3HMG 3-Hydroxy-3-methylglutarylcarnitin, C14_1 Tetradecenoylcarnitine, C14 Tetradecanoylcarnitine, C14OH Hydroxytetradecanoylcarnitin, C16_1 Hexadecenoylcarnitine, C16 Hexadecanoylcarnitine, C16_1OH Hydroxyhexadecenoylcarnitine, C16OH Hydroxyhexadecanoylcarnitine, C18_2 Octadecadienylcarnitine, C18_1 Octadecenoylcarnitine, C18 Octadecanoylcarnitine, C18_1OH Hydroxyoctadecenoylcarnitine, C18OH Hydroxyoctadecanoylcarnitin, Ala Alanine, Pro Proline, Val Valine, Thr Threonine, Leu/Ile Leucine / Isoleucine, Gln Glutamine, Met Methionine, His Histidine, Phe Phenylalanine, Tyr Tyrosine, Asp Aspartate, Glu Glutamate, Trp Tryptophan, Gly Glycine, Orn Ornithine, Arg Arginine, Cit Citrulline, HciHomocitrulline, Asa Argininosuccinat.

**Supplementary Table S3.** Metabolic and anthropometric characteristics of responders and non-responders.

| Parameter | Group | V0 | V3 | V6 | V7 |
|---|---|---|---|---|---|
| **Glycemia** | | | | | |
| FPG (mg/dl) | M-Diet Non-Resp. | 160.2±16.8 (11) | 145.9±20.1 (10) | 154.0±18.2 (10) | 178.1±29.6 (9) |
| | M-Diet Resp. | 156.0±11.2 (8) | 153.0±7.4 (8) | 146.5±13.0 (6) | 151.667±11.624 (6) |
| | FMD Non-Resp. | 164.9±14.3 (10) | 162.1±25.8 (9) | 154.3±23.9 (7) | 154±14.1 (6) |
| | FMD Resp. | 159.5±13.7 (11) | 145.9±11.2 (11) | 135.6±13.5 (10) | 150.7±11.1 (10) |
| HbA1c (%) | M-Diet Non-Resp. | 7.6±0.4 (11) | 7.0±0.4 (10) | 7.8±0.4 (10) | 8.1±0.6 (9) |
| | M-Diet Resp. | 7.7±0.4 (8) | 7.3±0.3 (8) | 7.7±0.4 (6) | 7.7±0.5 (6) |
| | FMD Non-Resp. | 8.5±0.6 (10) | 7.6±0.5 (9) | 6.7±0.6 (7) | 7.4±0.5 (6) |
| | FMD Resp. | 7.7±0.5 (11) | 7.0±0.3 (11) | 6.6±0.4 (10) | 7.7±0.6 (10) |
| C-Peptid (ng/ml) | M-Diet Non-Resp. | 3.5±0.6 (11) | 3.1±0.6 (10) | 3.5±0.7 (10) | 3.0±0.5 (8) |
| | M-Diet Resp. | 3.1±0.6 (8) | 2.7±0.6 (8) | 2.8±0.9 (6) | 2.9±0.9 (6) |
| | FMD Non-Resp. | 2.6±0.6 (10) | 2.6±0.8 (9) | 2.1±0.5 (7) | 2.5±0.8 (6) |
| | FMD Resp. | 3.5±0.4 (11) | 2.1±0.3 (11) | 2.0±0.3 (10) | 2.9±0.3 (9) |
| HOMA-IR | M-Diet Non-Resp. | 5.5±2.4 (7) | 5.0±2.7 (7) | 6.1±3.3 (6) | 6.9±4.1 (5) |
| | M-Diet Resp. | 5.8±1.1 (6) | 5.8±1.4 (6) | 5.8±2.0 (5) | 5.8±1.4 (5) |
| | FMD Non-Resp. | 5.0±1.8 (6) | 2.8±0.9 (6) | 2.7±1.4 (5) | 3.6±0.6 (4) |
| | FMD Resp. | 4.5±0.4 (7) | 1.8±0.3 (7) | 1.6±0.3 (6) | 3.6±1.9 (5) |
| **Blood pressure** | | | | | |
| Systolic BP | M-Diet Non-Resp. | 143.4±4.167 (10) | 146.7±9.107 (10) | 142.1±6.03 (10) | 138.3±4.6 (8) |
| | M-Diet Resp. | 140.3±7.0 (8) | 142±7.428 (8) | 143.167±5.18 (6) | 134.3±3.1 (6) |
| | FMD Non-Resp. | 146.4±3.8 (10) | 143.1±3.3 (8) | 147.0±3.7 (7) | 157.8±5.4 (6) |
| | FMD Resp. | 139.8±4.9 (11) | 130.6±4.4 (11) | 136.9±4.4 (9) | 162.8±2.8 (9) |
| Diastolic BP | M-Diet Non-Resp. | 82.6±2.0 (10) | 82.5±3.6 (10) | 84.9±3.3 (10) | 81.9±3.6 (8) |
| | M-Diet Resp. | 80.9±3.1 (8) | 79.9±1.7 (8) | 76.5±1.4 (6) | 78.7±2.3 (6) |
| | FMD Non-Resp. | 85.5±1.9 (10) | 84.6±2.3 (8) | 83.7±3.2 (7) | 85.8±4.1 (6) |
| | FMD | 85.2±3.0 | 79.5±1.8 | 82.8±2.2 | 85.4±3.3 (8) |

| | | | | | |
|---|---|---|---|---|---|
| | Resp. | (11) | (11) | (9) | |
| **Lipidemia** | | | | | |
| Cholesterol (mg/dl) | M-Diet Non-Resp. | 179.5±14.7 (11) | 174.2±12.7 (10) | 173.6±13.7 (10) | 171.0±15.7 (9) |
| | M-Diet Resp. | 168.4±15.1 (8) | 168.5±16.7 (8) | 201.8±18.4 (6) | 215.2±20.8 (6) |
| | FMD Non-Resp. | 197±35.6 (10) | 178.4±23.5 (9) | 161.9±10.8 (7) | 170.7±23.2 (6) |
| | FMD Resp. | 173.4±14.6 (11) | 153.5±14.2 (11) | 159±16.2 (10) | 182.1±13.8 (10) |
| LDL (mg/dl) | M-Diet Non-Resp. | 101.0±13.2 (10) | 96.6±8.8 (10) | 91.7±9.7 (10) | 94.5±13.6 (8) |
| | M-Diet Resp. | 86.0±11.7 (8) | 82.3±13.0 (8) | 104.7±16.4 (6) | 118.8±24.1 (5) |
| | FMD Non-Resp. | 90.5±13.3 (10) | 112.4±18.8 (8) | 88.6±8.4 (7) | 65.4±15.9 (5) |
| | FMD Resp. | 85.1±11.4 (10) | 82.7±13.9 (11) | 86.3±15.7 (10) | 95.3±12 (9) |
| HDL (mg/dl) | M-Diet Non-Resp. | 42.1±2.5 (11) | 41.2±1.9 (10) | 42.4±2.1 (10) | 41.7±4.0 (9) |
| | M-Diet Resp. | 44.3±5.1 (8) | 48.1±5.5 (8) | 54.0±4.8 (6) | 51.8±6.7 (6) |
| | FMD Non-Resp. | 38.5±2.8 (10) | 36.9±2.9 (9) | 43.3±4.7 (7) | 45.3±5.5 (6) |
| | FMD Resp. | 49.6±4.2 (11) | 48.5±3.5 (11) | 52.6±4.2 (10) | 55.5±4.9 (10) |
| TG (mg/dl) | M-Diet Non-Resp. | 216.0±41.5 (11) | 181.7±26.2 (10) | 197.8±25.6 (10) | 212.8±53.3 (9) |
| | M-Diet Resp. | 191.3±25.5 (8) | 190.3±34.4 (8) | 216.3±53.1 (6) | 217.5±43.3 (6) |
| | FMD Non-Resp. | 353.4±144.0 (10) | 236.3±77.8 (9) | 149.6±21.5 (7) | 245.2±59.9 (6) |
| | FMD Resp. | 208.0±62.0 (11) | 109.9±13.0 (11) | 101.3±11.9 (10) | 188.0±45.7 (10) |
| Lp(a) (mg/dl) | M-Diet Non-Resp. | 15.5±4.1 (11) | 17.8±5.8 (9) | 13.9±2.3 (10) | 13.7±2.1 (8) |
| | M-Diet Resp. | 50.8±18.0 (8) | 55.6±19.6 (8) | 66.3±19.8 (6) | 62.5±20.2 (6) |
| | FMD Non-Resp. | 20.7±7.2 (10) | 26.4±9.4 (9) | 19.4±3.8 (7) | 15.6±3.4 (6) |
| | FMD Resp. | 12.2±1.2 (11) | 15.9±2.3 (11) | 16.2±2.3 (10) | 13.4±1.7 (9) |
| **Liver parameters** | | | | | |
| AST (U/l) | M-Diet Non-Resp. | 27.5±2.3 (11) | 28.8±1.8 (10) | 26.7±1.5 (10) | 26.4±2.2 (9) |
| | M-Diet Resp. | 27.6±2.9 (8) | 33.9±6.6 (8) | 28.0±3.8 (6) | 26.2±1.8 (6) |
| | FMD Non-Resp. | 29.7±6.8 (10) | 33.3±4.7 (9) | 25.4±1.9 (7) | 26±2.1 (6) |
| | FMD Resp. | 25.5±1.6 (11) | 27.0±1.9 (11) | 25.6±2.0 (10) | 21.2±2.1 (10) |
| ALT (U/l) | M-Diet | 29.7±6.8 (10) | 33.3±4.7 (9) | 25.4±1.9 (7) | 29.6±3.6 (9) |
| | M-Diet Resp. | 25.5±1.6 (11) | 27.0±1.9 (11) | 25.6±2.0 (10) | 32.3±4.5 (6) |

| | | | | | |
|---|---|---|---|---|---|
| | FMD Non-Resp. | 30.5±5.9 (10) | 36.2±6.8 (9) | 26.0±3.0 (7) | 23.2±1.4 (6) |
| | FMD Resp. | 30.7±3 (11) | 33.5±3.2 (11) | 25.4±2.6 (10) | 23.4±3.8 (10) |
| Liver stiffness (kPa) | M-Diet Non-Resp. | 7.3±1.3 (11) | 6.2±0.9 (10) | 5.8±0.8 (10) | 7.9±2.0 (8) |
| | M-Diet Resp. | 6.5±0.7 (8) | 6.7±0.9 (8) | 6.4±1.1 (6) | 6.1±0.9 (6) |
| | FMD Non-Resp. | 10.5±1.9 (10) | 7.4±1.1 (8) | 8.4±0.8 (7) | 8.6±2.0 (6) |
| | FMD Resp. | 5.9±0.4 (11) | 5.4±0.5 (11) | 5.1±0.7 (10) | 6.1±0.4 (9) |
| **Protein Level** | | | | | |
| Plasma-Albumin (g/l) | M-Diet Non-Resp. | 43.7±1.2 (11) | 44.2±1.0 (10) | 44.2±0.8 (10) | 44.6±1.0 (9) |
| | M-Diet Resp. | 45.1±0.3 (8) | 45.6±0.4 (8) | 46.0±1.3 (6) | 44.7±0.8 (6) |
| | FMD Non-Resp. | 46.3±0.6 (10) | 46.5±1.2 (9) | 48.3±0.8 (7) | 46.1±0.6 (6) |
| | FMD Resp. | 45.5±0.6 (11) | 46.9±0.4 (11) | 46.6±0.7 (10) | 45.1±0.8 (10) |
| Plasma-total protein (g/l) | M-Diet Non-Resp. | 73.6±2.3 (11) | 72.8±2.5 (10) | 73.3±2.2 (10) | 73.0±2.0 (9) |
| | M-Diet Resp. | 72.1±0.8 (8) | 72.8±0.9 (8) | 71.0±1.5 (6) | 69.3±1.6 (6) |
| | FMD Non-Resp. | 74.2±1.4 (10) | 74.3±1.8 (9) | 71.7±0.9 (7) | 69.2±1.3 (6) |
| | FMD Resp. | 72.6±1 (11) | 74.8±1.3 (11) | 73.7±1.4 (10) | 69.7±1.0 (10) |
| **Renal parameters** | | | | | |
| **uACR (mg/g)** | M-Diet Non-Resp. | 112.1(302.6) (11) | 140.4(164.4) (10) | 275.6(360.8) (10) | 150.8(303.0) (9) |
| | M-Diet Resp. | **43.9(53.6) (8)*** | **23.1(46.4) (8)*** | **17.7(30.7) (6)*** | **19.7(11.4) (6)*** |
| | FMD Non-Resp. | 56.9(245.3) (10) | 92.0(384.3) (9) | 44.6(522.4) (7) | 25.2(676.4) (6) |
| | FMD Resp. | **51.3(104.1) (11)*** | **22.8(19.9) (11)*** | **23.4(23.9) (10)*** | **41.4(118.0) (10)*** |
| Urea (mg/dl) | M-Diet Non-Resp. | 39.9±5.2 (11) | 38.4±3.4 (10) | 51.4±13.5 (10) | 33.1±2.4 (9) |
| | M-Diet Resp. | 38.0±3.3 (8) | 37.6±5.4 (8) | 44.5±7.9 (6) | 40.0±4.2 (6) |
| | FMD Non-Resp. | 44.4±7.8 (10) | 26.6±3.4 (9) | 25.7±3.1 (7) | 38.5±3.2 (6) |
| | FMD Resp. | 33.6±2.4 (11) | 24.7±2.2 (11) | 26.6±2.9 (10) | 38±4.3 (10) |
| Creatinine (mg/dl) | M-Diet Non-Resp. | 0.8±0.1 (11) | 0.9±0.1 (10) | 0.9±0.1 (10) | 0.8±0.1 (9) |
| | M-Diet Resp. | 0.8±0.1 (8) | 0.8±0.1 (8) | 0.9±0.1 (6) | 0.8±0.1 (6) |
| | FMD Non-Resp. | 0.9±0.1 (10) | 0.9±0.1 (9) | 0.9±0.1 (7) | 0.9±0.1 (6) |
| | FMD Resp. | 0.9±0.1 (11) | 0.9±0.1 (11) | 0.9±0.1 (10) | 0.9±0.1 (10) |
| eGFR CKD-EPI(creatinine) | M-Diet Non-Resp. | 87.7±6.5 (11) | 84.7±6.7 (10) | 85.4±6.0 (10) | 87.2±5.8 (9) |

| | | | | | |
|---|---|---|---|---|---|
| (ml/min/1,73m²) | M-Diet Resp. | 86.0±5.6 (8) | 85.4±6.1 (8) | 88.1±8.1 (6) | 88.4±7.0 (6) |
| | FMD Non-Resp. | 87.5±5.8 (10) | 90±5 (9) | 85.1±6.3 (7) | 85.1±6.2 (6) |
| | FMD Resp. | 81.9±6.4 (11) | 77.7±6.1 (11) | 79.4±7 (10) | 80.6±6.7 (10) |
| Cystatin C (mg/l) | M-Diet Non-Resp. | 1.0±0.1 (11) | 1.1±0.1 (10) | 1.2±0.1 (10) | 1.1±0.1 (9) |
| | M-Diet Resp. | 1.0±0.1 (8) | 1.0±0.1 (8) | 1.1±0.1 (6) | 1.1±0.1 (6) |
| | FMD Non-Resp. | 1.1±0.2 (10) | 1.1±0.1 (9) | 1.0±0.1 (7) | 1.1±0.1 (6) |
| | FMD Resp. | 1.1±0.1 (11) | 1.1±0.1 (11) | 1.1±0.1 (10) | 1.1±0.1 (10) |
| eGFR (Cystatin C) (ml/min/1,73m²) | M-Diet Non-Resp. | 81.6±6.7 (11) | 76.7±6.2 (10) | 73.3±8.2 (10) | 76.8±6.8 (9) |
| | M-Diet Resp. | 88.7±5.7 (8) | 84.1±5.7 (8) | 72.3±7.1 (6) | 68.0±7.0 (6) |
| | FMD Non-Resp. | 82.4±7.0 (10) | 81.0±7.7 (9) | 79.7±7.9 (7) | 70.2±7.9 (6) |
| | FMD Resp. | 76.8±6.4 (11) | 74.1±5.4 (11) | 75.7±6.9 (10) | 71.7±6.6 (10) |
| sUPAR (pg/ml) | M-Diet Non-Resp. | 2908.1±202.6 (11) | 3463.1±607.3 (9) | 3074.4±277.2 (10) | 3269.6±307.7 (8) |
| | M-Diet Resp. | 3222.5±448.9 (8) | 2704.3±170.9 (8) | 2810.2±205.3 (6) | 2763.8±171.9 (6) |
| | FMD Non-Resp. | 3542.9±765 (10) | 3233.5±444.2 (9) | 2764.5±146.7 (7) | 3128.6±327.6 (6) |
| | FMD Resp. | 2739.3±165.4 (11) | 2806.3±158.6 (11) | 2717.9±178.3 (10) | 2975.5±221.4 (9) |
| **Ketogenesis** | | | | | |
| Blood ketones (mmol/l) | M-Diet Non-Resp. | 0.2±0.0 (11) | 0.2±0.1 (10) | 0.1±0.0 (10) | 0.1±0.0 (8) |
| | M-Diet Resp. | 0.1±0.0 (8) | 0.1±0.0 (8) | 0.1±0.0 (6) | 0.1±0.0 (6) |
| | FMD Non-Resp. | 0.2±0.1 (10) | 0.6±0.2 (9) | 0.6±0.2 (7) | 0.1±0.0 (6) |
| | FMD Resp. | 0.1±0 (11) | 0.4±0.1 (11) | 0.4±0.1 (10) | 0.1±0.0 (10) |
| Urine ketones (mg/dl) | M-Diet Non-Resp. | 0.0±0.0 (11) | 3.0±1.5 (10) | 2.0±1.5 (10) | 1.3±0.8 (8) |
| | M-Diet Resp. | 0.6±0.6 (8) | 1.3±0.8 (8) | 3.3±2.5 (6) | 2.5±2.5 (6) |
| | FMD Non-Resp. | 2.0±1.5 (10) | 6.7±2.2 (9) | 7.9±5.8 (7) | 1.7±1.1 (6) |
| | FMD Resp. | 0.5±0.5 (11) | 5.0±2 (11) | 8.0±3.8 (10) | 0.6±0.6 (9) |
| **Other metabolic parameters** | | | | | |
| Uric acid (mg/dl) | M-Diet Non-Resp. | 6.2±0.5 (11) | 6.3±0.5 (10) | 6.3±0.6 (10) | 6.0±0.4 (8) |
| | M-Diet Resp. | 5.7±0.4 (8) | 5.8±0.4 (8) | 5.5±0.5 (6) | 5.8±0.6 (6) |
| | FMD Non-Resp. | 6.6±0.5 (10) | 6.4±0.4 (9) | 6.8±0.4 (7) | 6.3±0.4 (6) |
| | FMD Resp. | 6.6±0.5 (11) | 6.4±0.5 (11) | 6.8±0.6 (10) | 6.4±0.8 (9) |

| | | | | | |
|---|---|---|---|---|---|
| hsCRP (mg/l) | M-Diet Non-Resp. | 1.8±0.6 (11) | 2.0±0.7 (10) | 1.8±0.5 (10) | 3.3±1.5 (9) |
| | M-Diet Resp. | 2.0±0.6 (8) | 2.2±0.8 (8) | 2.9±1.2 (6) | 3.0±1.3 (6) |
| | FMD Non-Resp. | 4.0±1.9 (10) | 5.1±2.1 (9) | 4.4±3.0 (7) | 3.5±2.5 (6) |
| | FMD Resp. | 6.1±2.6 (11) | 1.6±0.3 (11) | 1.4±0.3 (10) | 1.7±0.5 (10) |
| hsTNT (pg/ml) | M-Diet Non-Resp. | 16.6±2.8 (11) | 18.0±3.4 (10) | 19.0±4.3 (10) | 17.4±3.8 (9) |
| | M-Diet Resp. | 14.9±3.6 (8) | 15.8±3.6 (8) | 17.2±6.7 (6) | 16.7±5.8 (6) |
| | FMD Non-Resp. | 15.2±2.7 (10) | 14.2±3.3 (9) | 14.3±2.9 (7) | 23.5±5.8 (6) |
| | FMD Resp. | 10.3±0.9 (11) | 11.5±1.2 (11) | 12.1±1.1 (10) | 11.4±1.3 (10) |
| IGF-1 (ng/ml) | M-Diet Non-Resp. | 95.8±10.6 (11) | 102.1±13.1 (10) | 102.8±12.3 (10) | 117.4±17.4 (8) |
| | M-Diet Resp. | 134.1±9.8 (8) | 136.2±12.5 (8) | 135.6±16.3 (6) | 132.0±19.4 (6) |
| | FMD Non-Resp. | 130±11.0 (10) | 133.2±11.7 (9) | 133.7±16.3 (7) | 125.7±12.8 (6) |
| | FMD Resp. | 139.6±13.5 (11) | 123.3±8.7 (11) | 123.3±10.8 (9) | 144.9±9.3 (9) |
| IL-6 (pg/ml) | M-Diet Non-Resp. | 3.7±1.1 (11) | 4.0±0.7 (10) | 4.1±1.3 (9) | 3.4±0.6 (8) |
| | M-Diet Resp. | 2.0±0.1 (8) | 3.5±1.1 (8) | 1.9±0.0 (6) | 2.2±0.2 (6) |
| | FMD Non-Resp. | 3.1±0.9 (10) | 3.8±1.2 (9) | 2.3±0.2 (7) | 2.6±0.4 (6) |
| | FMD Resp. | 2.6±0.3 (11) | 3.3±0.6 (11) | 2.2±0.2 (10) | 2.0±0.0 (8) |
| NT-proBNP (ng/l) | M-Diet Non-Resp. | 122.1±29.1 (11) | 174.8±42.1 (9) | 129.0±33.0 (10) | 135.4±31.4 (9) |
| | M-Diet Resp. | 187.6±85.6 (8) | 163.0±57.9 (8) | 129.0±70.9 (6) | 147.8±98.1 (6) |
| | FMD Non-Resp. | 296.8±209.6 (10) | 258.8±138.3 (9) | 218.4±129.1 (7) | 430.3±282.2 (6) |
| | FMD Resp. | 109.1±27.6 (11) | 151.3±51.9 (10) | 142.1±43.7 (10) | 146.3±55.5 (10) |
| **Body weight** | | | | | |
| Body weight (kg) | M-Diet Non-Resp. | 96.2±4.8 (11) | 95.9±5.3 (10) | 95.7±5.539 (10) | 90.4±5.6 (8) |
| | M-Diet Resp. | 89.3±5.4 (8) | 89.3±5.9 (8) | 88.8±7.4 (6) | 90.6±8.0 (6) |
| | FMD Non-Resp. | 102.8±5.7 (10) | 98.9±6.1 (9) | 94.2±5.9 (7) | 99.8±6.6 (6) |
| | FMD Resp. | 99.2±3.8 (11) | 92.7±3.6 (11) | 89.9±3.6 (10) | 98.9±3.6 (10) |
| BMI ($kg/m^2$) | M-Diet Non-Resp. | 30.8±1.1 (11) | 30.5±1.2 (10) | 30.6±1.3 (10) | 29.6±1.3 (8) |
| | M-Diet Resp. | 29.4±1.8 (8) | 29.5±2.1 (8) | 29.5±2.6 (6) | 30.1±2.7 (6) |
| | FMD Non-Resp. | 31.8±1.5 (10) | 29.7±1.8 (9) | 28.2±1.9 (7) | 30.8±2.2 (6) |
| | FMD Resp. | 29.2±1 (11) | 27.0±1.0 (11) | 26.0±0.9 (10) | 27.7±1.0 (10) |

| | | | | | |
|---|---|---|---|---|---|
| **Body composition (BIA)** | | | | | |
| Fat mass (%) | M-Diet Non-Resp. | 30.0±2.2 (10) | 30.0±2.2 (9) | 30.2±2.3 (9) | 72.0±3.4 (8) |
| | M-Diet Resp. | 29.8±3.6 (8) | 30.1±3.4 (8) | 30.3±4.2 (6) | 68.3±4.2 (6) |
| | FMD Non-Resp. | 32.0±2.3 (10) | 31.7±3.2 (9) | 29.9±3.8 (6) | 29.1±3.9 (6) |
| | FMD Resp. | 31.9±2.7 (11) | 30.7±2.4 (11) | 29.3±2.5 (10) | 30.7±2.8 (9) |
| Fat mass (kg) | M-Diet Non-Resp. | 28.7±2.8 (10) | 28.7±3.2 (9) | 28.6±3.1 (9) | 64.3±4.8 (8) |
| | M-Diet Resp. | 28.1±5.2 (8) | 28.0±4.9 (8) | 28.3±6.1 (6) | 60.5±3.2 (6) |
| | FMD Non-Resp. | 33.1±3.2 (10) | 31.9±4.3 (9) | 28.1±4.5 (6) | 29.3±4.6 (6) |
| | FMD Resp. | 28.6±2.8 (11) | 25.6±2.4 (11) | 23.6±2.4 (10) | 26.6±2.8 (9) |
| Fat free mass (%) | M-Diet Non-Resp. | 70.0±2.2 (10) | 70.0±2.2 (9) | 69.7±2.3 (9) | 26.3±2.6 (8) |
| | M-Diet Resp. | 70.2±3.6 (8) | 69.9±3.4 (8) | 69.7±4.2 (6) | 30.1±6.3 (6) |
| | FMD Non-Resp. | 68±2.3 (10) | 68.3±3.2 (9) | 70.2±3.8 (6) | 71.0±3.9 (6) |
| | FMD Resp. | 68.1±2.7 (11) | 69.3±2.4 (11) | 70.7±2.5 (10) | 69.3±2.8 (9) |
| Fatt free mass (kg) | M-Diet Non-Resp. | 66.7±3.8 (10) | 66.1±4.0 (9) | 65.9±4.4 (9) | 72.0±3.4 (8) |
| | M-Diet Resp. | 62.4±2.5 (8) | 61.2±2.2 (8) | 60.5±2.4 (6) | 68.3±4.2 (6) |
| | FMD Non-Resp. | 69.8±4.3 (10) | 67.0±4.3 (9) | 64.7±4.8 (6) | 70.5±5.5 (6) |
| | FMD Resp. | 60.4±3.1 (11) | 57.1±2.6 (11) | 56.3±3.0 (10) | 59.2±2.9 (9) |
| Body liquid (%) | M-Diet Non-Resp. | 53.3±1.7 (10) | 53.2±1.9 (9) | 52.5±1.8 (9) | 53.2±1.9 (8) |
| | M-Diet Resp. | 53.2±2.9 (8) | 52.5±2.5 (8) | 52.6±3.3 (6) | 51.4±3.4 (6) |
| | FMD Non-Resp. | 51.5±2 (10) | 51.5±2.5 (9) | 51.9±2.5 (6) | 54.1±2.5 (6) |
| | FMD Resp. | 50.5±2.1 (11) | 50.9±1.7 (11) | 51.5±1.9 (10) | 51.8±2.0 (9) |
| Body liquid (l) | M-Diet Non-Resp. | 50.8±3.0 (10) | 50.3±3.1 (9) | 49.6±3.5 (9) | 48.5±3.8 (8) |
| | M-Diet Resp. | 47.2±2.1 (8) | 45.9±1.5 (8) | 45.65±1.93 (6) | 45.5±2.5 (6) |
| | FMD Non-Resp. | 53.1±3.8 (10) | 50.9±3.9 (9) | 48.0±4.0 (6) | 53.9±4.3 (6) |
| | FMD Resp. | 44.9±2.6 (11) | 42±2.1 (11) | 41.2±2.6 (10) | 44.4±2.4 (9) |
| Phase angel | M-Diet Non-Resp. | 5.9±0.2 (10) | 5.9±0.2 (9) | 6.0±0.2 (9) | 5.8±0.2 (8) |
| | M-Diet Resp. | 5.7±0.2 (8) | 5.7±0.2 (8) | 5.6±0.2 (6) | 5.3±0.2 (6) |
| | FMD Non-Resp. | 6.0±0.2 (10) | 5.7±0.3 (9) | 6.2±0.1 (6) | 5.8±0.3 (6) |
| | FMD Resp. | 5.7±0.3 (11) | 5.6±0.2 (11) | 5.5±0.2 (10) | 5.4±0.3 (9) |

Data are shown as mean ± SEM of unadjusted values of variable for normally distributed variables or as median (IQR) for log-normally distributed variables. Patients in both groups that showed a decrease in albuminuria of at least 30% compared to the respective baseline are referred as Responders, while the rest are referred as non-Responders. Comparison of responders vs. non-responders is done by ANCOVA with adjustment for age, sex and weight loss. *P≤0.05.

Abbreviations: M-Diet Mediterranean diet, FMD fasting-mimicking diet, FPG fasting plasma glucose, HOMA-IR Homeostatic Model Assessment of Insulin Resistance, LDL low-density lipoprotein, HDL high-density lipoprotein, TG triglyceride, Lp(a) lipoprotein (a), ALT alanine aminotransferase, AST aspartate aminotransferase, uACR urinary albumin-to-creatinine ratio, Micro.uACR microalbuminuria, Macro.uACR macroalbuminuria, eGFR estimated glomerular filtration rate, sUPAR soluble urokinase-type plasminogen activator receptor, hsCRP high-sensitivity C-reactive protein, hsTNT high-sensitivity troponin T, IGF-1 insulin-like growth factor 1, IL-6 interleukin 6, NT-proBNP N-terminal prohormone of brain natriuretic peptide, BMI body mass index.

**Supplementary Table S4**. Metabolic and anthropometric parameters at baseline and at refeeding after 3 (rV3) and after 6 diet cycles (rV6).

| **Parameter** | **Group** | **V0** | **rV3** | **rV6** |
|---|---|---|---|---|
| **Glycemia** | | | | |
| FPG (mg/dl) | M-Diet | 158.4±10.6 (19) | 147.8±12.5 (16) | 147.5±11.0 (16) |
| | FMD | 162.1±9.7 (21) | 149.8±11.4 (20) | 148.9±7.8 (17) |
| HbA1c (%) | M-Diet | 7.7±0.3 (19) | 7.2±0.3 (16) | 7.5±0.3 (16) |
| | FMD | 8.1±0.4 (21) | 7.2±0.3 (20) | 6.9±0.3 (17) |
| C-Peptid (ng/ml) | M-Diet | 3.3±0.4 (19) | 2.9±0.5 (16) | 2.9±0.4 (16) |
| | FMD | 3.1±0.4 (21) | 3.3±0.4 (20) | 3.3±0.4 (17) |
| HOMA-IR | M-Diet | 6.1±0.9 (13) | 5.2±0.8 (11) | 5.5±0.9 (11) |
| | FMD | 6.4±1.9 (14) | 5.6±1.5 (14) | 6.3±1.9 (12) |
| **Blood pressure** | | | | |
| Systolic BP | M-Diet | 142.0±3.8 (19) | N.A. | N.A. |
| | FMD | 142.9±3.1 (21) | N.A. | N.A. |
| Diastolic BP | M-Diet | 81.8±1.7 (19) | N.A. | N.A. |
| | FMD | 85.3±1.8 (21) | N.A. | N.A. |
| **Lipidemia** | | | | |
| Cholesterol (mg/dl) | M-Diet | 174.8±10.4 (19) | 168.2±10.2 (16) | 181.2±10.2 (16) |
| | FMD | 184.6±18.3 (21) | 172.4±13.5 (20) | 172.9±10.4 (17) |
| LDL (mg/dl) | M-Diet | 94.3±8.9 (18) | 88.6±7.7 (16) | 93.6±7.8 (16) |
| | FMD | 87.8±8.5 (20) | 81.6±8.5 (18) | 85.9±9.4 (16) |
| HDL (mg/dl) | M-Diet | 43±2.5 (19) | 44.2±3.3 (16) | 47.7±3 (16) |
| | FMD | 44.3±2.8 (21) | 45.6±2.6 (20) | 53.8±3.8 (17) |
| TG (mg/dl) | M-Diet | 205.6±25.8 (19) | 177.1±20.0 (16) | 199.8±24.8 (16) |
| | FMD | 277.2±75.6 (21) | 241.2±50.2 (20) | 179.3±27.5 (17) |
| Lp(a) (mg/dl) | M-Diet | 30.3±8.7 (19) | N.A. | N.A. |
| | FMD | 16.3±3.5 (21) | N.A. | N.A. |
| **Liver parameters** | | | | |
| AST (U/l) | M-Diet | 27.5±1.7 (19) | 29.6±2.2 (16) | 28.2±2 (16) |
| | FMD | 27.5±3.3 (21) | 23.4±1.9 (20) | 22.2±1.4 (17) |
| ALT (U/l) | M-Diet | 31.3±2.8 (19) | 31.6±3.3 (16) | 30.4±3.3 (16) |
| | FMD | 30.6±3.1 (21) | 25.9±2.2 (20) | 23.5±1.7 (17) |
| Liver stiffness(kPa) | M-Diet | 6.9±0.8 (19) | N.A. | N.A. |
| | FMD | 8.1±1 (21) | N.A. | N.A. |
| **Protein Level** | | | | |
| Plasma-Albumin (g/l) | M-Diet | 44.3±0.7 (19) | 43.8±0.9 (16) | 44.8±0.8 (16) |
| | FMD | 45.9±0.4 (21) | 45.5±0.6 (20) | 45.9±0.5 (17) |
| Plasma-total protein (g/l) | M-Diet | 72.9±1.4 (19) | 71.4±1.5 (16) | 71.5±1.4 (16) |
| | FMD | 73.3±0.9 (21) | 72.1±1.2 (20) | 70.5±0.9 (17) |

| | | | | |
|---|---|---|---|---|
| **Renal parameters** | | | | |
| uACR (mg/g) | M-Diet | 73.4(205.6) (19) | 36.5(111.9) (16) | 63.8(306.7) (16) |
| | FMD | 51.3(116.0) (21) | 45.1(123.6) (20) | 42.2(98.4) (17) |
| Micro.uACR (mg/g) | M-Diet | 44.8(49.2) (14) | 23.0(39.6) (12) | 22.9(65.6) (11) |
| | FMD | 43.7(39.6) (17) | 29.5(55.6) (16) | 27.4(27.1) (14) |
| Macro.uACR (mg/g) | M-Diet | 359.3(1888.5) (4) | 323.4(560.0) (4) | 399.8(143.4) (5) |
| | FMD | 555.2(593.2) (3) | 714.8(593.0) (4) | 915.4(905.8) (3) |
| Urea (mg/dl) | M-Diet | 39.1±3.2 (19) | 38.4±4.5 (16) | 40.1±3.4 (16) |
| | FMD | 38.8±4.0 (21) | 34.2±2.5 (20) | 39.0±5.4 (17) |
| Creatinine (mg/dl) | M-Diet | 0.8±0.1 (19) | 0.9±0.1 (16) | 0.8±0.1 (16) |
| | FMD | 0.9±0.1 (21) | 0.9±0.1 (20) | 0.8±0.1 (17) |
| eGFR CKD-EPI(creatinine) (ml/min/1,73m²) | M-Diet | 87.0±4.3 (19) | 84.7±4.3 (16) | 86.5±4.5 (16) |
| | FMD | 84.6±4.3 (21) | 85.1±4.1 (20) | 86.5±4.4 (17) |
| Cystatin C (mg/l) | M-Diet | 1.0±0.1 (19) | 1.0±0.1 (16) | 1.1±0.1 (16) |
| | FMD | 1.1±0.1 (21) | 1.1±0.1 (20) | 1.1±0.1 (17) |
| eGFR (Cystatin C) (ml/min/1,73m²) | M-Diet | 84.6±4.5 (19) | 83.5±4.5 (16) | 73.6±5.1 (16) |
| | FMD | 79.5±4.6 (21) | 76.4±4.6 (20) | 75.2±5.1 (17) |
| sUPAR (pg/ml) | M-Diet | 3040.5±218.0 (19) | N.A. | N.A. |
| | FMD | 3122.0±375.0 (21) | N.A. | N.A. |
| **Ketogenesis** | | | | |
| Blood ketones (mmol/l) | M-Diet | 0.1±0.0 (19) | 0.1±0.0 (16) | 0.1±0.0 (16) |
| | FMD | 0.2±0.0 (21) | 0.2±0.0 (20) | 0.1±0.0 (17) |
| Urine ketones (mg/dl) | M-Diet | 0.3±0.3 (19) | 3.1±1.3 (16) | 0.9±0.5 (16) |
| | FMD | 1.2±0.8 (21) | 1.5±0.8 (20) | 0.6±0.4 (17) |
| **Other metabolic parameters** | | | | |
| **Uric acid (mg/dl)** | M-Diet | 6.0±0.3 (19) | 6.1±0.4 (16) | 5.9±0.4 (16) |
| | FMD | 6.6±0.4 (21) | **5.8±0.3 (20)*** | 5.8±0.3 (17) |
| **hsCRP (mg/l)** | M-Diet | 1.9±0.4 (19) | 1.6±0.4 (16) | 6.5±3.8 (16) |
| | FMD | 5.1±1.6 (21) | 3.0±1.0 (20) | **2.7±0.9 (17)*** |
| hsTNT (pg/ml) | M-Diet | 15.9±2.2 (19) | 16.1±2.5 (16) | 18.6±3.1 (16) |
| | FMD | 12.6±1.5 (21) | 12.4±1.3 (20) | 13.9±1.9 (17) |
| IGF-1 (ng/ml) | M-Diet | 111.9±8.5 (19) | 119.7±10.2 (16) | 112.7±10.1 (16) |
| | FMD | 135.1±8.7 (21) | 141.3±8.4 (20) | 153.5±9.6 (17) |
| IL-6 (pg/ml) | M-Diet | 3.0±0.6 (19) | 3.8±0.6 (16) | 3.0±0.5 (16) |
| | FMD | 2.9±0.4 (21) | 3.2±0.6 (20) | 2.8±0.4 (17) |
| NT-proBNP (ng/l) | M-Diet | 149.7±39.1 (19) | N.A. | N.A. |
| | FMD | 198.5±100.3 (21) | N.A. | N.A. |
| Plasma MG-H1, nM | M-Diet | 217.9±36.0 (19) | 190.3± 29.3 (16) | 211.4±19.8 (16) |
| | FMD | 213.3±32.0 (21) | 168.0± 29.1 (20) | 189,8±38,7 (17) |
| **Body weight** | | | | |
| **Body weight (kg)** | M-Diet | 93.3±3.6 (19) | 90.3±3.7 (16) | 95.1±4.6 (15) |
| | FMD | 95.7±3.6 (21) | **91.7±3.9 (20)***** | **87.9±3.6 (17)*** |

| | | | | |
|---|---|---|---|---|
| **BMI (kg/m²)** | M-Diet | 30.2±1.0 (19) | 29.3±1.0 (16) | 30.6±1.3 (15) |
| | FMD | 31±0.9 (21) | **29.7±1.1 (20)**** | **28.4±1.0 (17)*** |

Data are shown as mean ± SEM of unadjusted values of variable for normally distributed variables or as median (IQR) for log-normally distributed variables. Statistical significant values are written in bold and indicate significance level of intervention effect on change of parameter compared to baseline and are based on M-Diet corrected ANCOVA with adjustment for age, sex and weight loss. *** P≤0.001, **P≤0.01, *P≤0.05.

*Abbreviations:* M-Diet Mediterranean diet, FMD fasting-mimicking diet, FPG fasting plasma glucose, HOMA-IR Homeostatic Model Assessment of Insulin Resistance, BP, blood pressure, TG triglyceride, Lp(a) lipoprotein (a), ALT alanine aminotransferase, AST aspartate aminotransferase, uACR urinary albumin-to-creatinine ratio, Micro.uACR microalbuminuria, Macro.uACR macroalbuminuria, eGFR estimated glomerular filtration rate, sUPAR soluble urokinase-type plasminogen activator receptor, hsCRP high-sensitivity C-reactive protein, hsTNT high-sensitivity troponin T, IL-6 interleukin 6, NT-proBNP N-terminal prohormone of brain natriuretic peptide.

#### SUPPLEMENTARY DATA

##### Inclusion criteria

To be considered eligible to participate in this study, participants had to meet all of the inclusion criteria listed below:

- Capacity to consent
- Obtained written informed consent
- Age between 50 and 75 years
- Diabetes duration at least 1 year
- Increased albuminuria levels (albuminuria concentration of ≥ 20 mg/l oder ≥ 20 mg/g urine-creatinine for men and ≥ 30 mg/g urine-creatinine for women) in two consecutive spot urine samples.
- eGFR>30 ml/min/173cm² from CKD-EPI formula
- Oral antidiabetic or insulin therapy:
- BMI 23-40 kg/m²

##### Exclusion criteria

In addition, to be eligible for enrolment in the trial, participants must not meet any of the exclusion criteria listed below:

- Legally incapacitated persons
- Other form of diabetes (i.e. diabetes mellitus type 1, pancreatogenic diabetes or steroid-induced diabetes)
- Acute infection/fever
- Immunsuppressive Therapy
- Severe heart, kidney or liver disease:
- NYHA Class IV
- Non-dibetic liver disease (i.e. primary bliliary cirrhosis, primary sclerosing cholangitis, Morbus Wilson, hemochromatosis, autoimmunehepatitis)
- Severe peripheral artery disease (stage IV)
- Non-diabetic glomerulopathy
- Alcohol or drug abuse
- History of cancer disease in the last 5 years prior to study
- Infectious Hepatatis B, C, E, HIV infection
- Autoimmune diseases or immunesuppressive therapy
- Participation in other interventional studies
- Anemia or hematological disease
- Other causes of polyneuropathy (autoimmune, alcohol-induced or Vitamin B12 deficiency, collagenosis)
- Pacemaker
- Food allergy (nuts, tomato, soja or other ingredients enlisted in the diet programm)

#### Table of assessments

| | Screen | Baseline | diet intervention period | | | | | | | | Follow-up |
|---|---|---|---|---|---|---|---|---|---|---|---|
| | | | 1st cycle | 2nd cycle | 3rd cycle | re-feeding | 4th cycle | 5th cycle | $6^{th}$ cycle | re-feeding | |
| **Visit** | **-V1** | **V0** | **V1** | **V2** | **V3** | **rV3** | **V4** | **V5** | **V6** | **rV6** | **V7** |
| **Study month** | **≤ 4 wks** | **≤ 2 wks** | **1 m** | **2 m** | **3 m** | **3 m** | **4 m** | **5 m** | **6** | **6 m** | **112 ± 31d** |
| Informed consent | | x | | | | | | | | | |
| Randomization | | x | | | | | | | | | |
| Demography and smoking | x | | | | | | | | | | |
| Medical history | x | | | | | | | | | | |
| Physical examination | x | x | | | x | | | | x | | x |
| Anthropometry | x | x | x | x | x | x | x | x | x | x | x |
| 12-lead ECG | | x | | | x | | | | x | | x |
| Bioimpedance analysis | | x | | | x | | | | x | | x |
| Diet counseling | x | x | | | x | x | | | x | x | |
| Assessment of changes in diet or excercise | | | x | x | x | x | x | x | x | x | |
| Blood chemistry, lipids, liver parameters renal function, glycemia, urine analysis | x | x | x | x | x | x | x | x | x | x | x |
| Ketone bodies | | x | x | x | x | x | x | x | x | x | x |
| MG-H1 | | x | | | x | x | | | x | x | x |
| AC profile | | x | | | x | x | | | x | x | x |
| MG Glyoxalase 1 | | x | | | x | x | | | | | |

| | | | | | | | | | | | |
|---|---|---|---|---|---|---|---|---|---|---|---|
| activity, yH2Ax, comet assay | | | | | | | | | | | |
| Hydroxyac-etone | | **x** | | | **x** | **x** | | | **x** | **x** | **x** |
| Assess. adverse events | | | **x** | **x** | **x** | **x** | **x** | **x** | **x** | **x** | |

**Quantification of acylcarnitines**

Acylcarnitines (AC) were determined in serum by electrospray ionization tandem mass spectrometry (ESI-MS/MS) according to a modified method as previously described (1), using a Quattro Ultima triple quadrupole mass spectrometer (Micromass, Manchester, UK) equipped with an electrospray ion source and a Micromass MassLynx data system. In particular, 5 µl of plasma were placed on a 4.7 mm filter paper punch, dried at room temperature overnight and extracted with 100 µl of deuterium-labeled standard solution in methanol (1).

**Measurement of MG, MG-H1, hydroxyacetone and glyoxalase 1 activity**

Plasma concentration of MG and plasma and urine concentration of MG-H1 was determined by stable isotopic dilution, LC-MS/MS, as described previously (2). Hydroxyacetone was determined using O-(2,3,4,5,6-pentafluorophenyl) methyl hydroxylamine (PFBHA) as a derivatizing agent (3). Activity of Glo1 was determined spectrophotometrically as described previously (3).

**Phosphorylated glyoxalase-1**

Total protein was isolated from white blood cells using IP buffer and protein concentration was determined using the Bradford reagent (4). Proteins were resolved by SDS-PAGE (15% Mini-PROTEAN TGX, Biorad) and transferred onto a 0.2 µM nitrocellulose membrane and blocked using 5% milk powder in PBS and 0.05% Tween 20 (PBS-T) at room temperature for 1hr. Membranes were then incubated overnight at 4 °C with antibodies against phosphorylated Y136 Glo1 Rabbit polyclonal self-made (1:250 dilution), Glo1 antibody ginea pig polyclonal self-made (1:1000) and Calnexin Rabbit polyclonal antibody Enzo (1:1000 dilution) in 5% BSA in PBS-T. Blots were washed and then incubated with an appropriate HRP-conjugated antibody (1:5000 in 5% milk powder in PBS-T) for 1hr at room temperature. Proteins were visualized on a ChemiDoc system using Western Lightning Plus-ECL (PerkinElmer) with varying exposure times (0.1–3 min). Protein expression was determined using the software Image-master (Bio-Rad) and normalized to calnexin.

**Isolation of white blood cells (WBC)**

Blood samples were processed immediately after taking the blood in fasting conditions as described above. All following steps were done on ice at 4°C to avoid sample degradation. One syringe of 9 ml EDTA blood (Sarstedt AG & Co. KG, Nümbrecht, Germany) was spun down at 3000 rpm for 5 min and 2 ml of plasma supernatant were transferred in a separate tube and spun again at 14.000rpm for 1 min. These plasma samples were frozen at -80°C until they were used for further analyses. One syringe of 9 ml EDTA blood (Sarstedt AG & Co. KG, Nümbrecht, Germany) was lysed in 40 ml erythrocyte lysis buffer (ECL, 1.5M ammonium chloride, 100mM sodium bicarbonate, 13mM EDTA pH7.3 and before use is diluted 1:10) for 15 min. Cells were spun down at 1700 rpm for 5 min. Afterwards the cells were washed once with ECL and once with NaCl 0.9% and spun down at 1300 rpm for 5 min. The cells were resuspended in fetal calve serum (FCS, Sigma-Aldrich, St. Louis, MO, USA) and frozen in FCS with 10 % DMSO (~3 Mio cells per tube) at -80°C until the samples were used for further analyses.

**Determination of Comet tail length and yH2Ax-positivity in WBC**

For each patient one vial with ~3 million cells were thawed in a 37 °C waterbath and immediately transferred into 10 ml prewarmed PBS (Sigma-Aldrich, St. Louis, MO, USA) and spun down at 1400 rpm for 10 min. Following a second wash-step with 10 ml PBS, cells were incubated with 1 ml of 200 ng DNase (Sigma-Aldrich, St. Louis, MO, USA) in PBS and 5 mM MgCl for 30 min at room temperature and spun down again. After resuspension in 1 ml PBS with 10 % FCS and 1 mM EDTA (FACS buffer), 10 µl per patient were transferred to a v-shaped 96 well plate for the Comet Assay (Trevigen, Gaithersburg, MD, USA). The remaining material was spun down, resuspended in 200 µl FACS buffer and 180 µl transferred to another v-shaped 96 well plate (Sigma-Aldrich, St. Louis, MO, USA) for the FACS analysis of yH2Ax. Remaining 20 µl per sample were pooled and used for controls.

**Comet -Assay**

Comet Assay was performed with the Reagent Kit for Higher Throughput Single Cell Gel Electrophoresis Assay (4252-040-K, Trevigen; Gaithersburg, MD, USA). The assay was performed according to the protocol, but with the following changes: For each sample 10 µl of cells were resuspended with 100 µl LMAgarose and 15 µl were spread on the comet plate. To adjust for the used electrophoresis apparatus, the alternative alkaline unwinding and electrophoresis solutions were used. Electrophoresis was run with 380 ml at 15 V for 1 hour in an ice bath. Comets were stained with SYBRGreen 1:10,000 (S7563, ThermoFisherScientific, Waltham, MA, USA). Plates were washed twice after staining. Pictures were taken with Olympus IX81 Widefield microscope using Olympus ScanR acquisition and comets were analysed with OpenComet (ImageJ). Mean-TailDNA% was used as readout.

**yH2Ax-positivity-measurements**

Plate was spun at 1300 rpm for 5 min and liquid discarded. Cells were incubated with 20 µl Fc Receptor Blocking Solution (422302, Biolegend, San Diego, CA, USA) 1:200 in FACS buffer for 10 min and after removal with 20 µl CD-antibodies (APC/Cyanine7 anti-human CD3 Antibody, 300425; Alexa Fluor® 700 anti-human CD4 Antibody, 317425; Brilliant Violet 421™ anti-human CD14 Antibody, 325628; APC anti-human CD19 Antibody, 302211, all Biolegend, San Diego, CA, USA) 1:100 in FACS buffer for

30 min. After washing with 100 µl FACS buffer, cells were fixed with 100 µl fixation buffer (420801, Biolegend, San Diego, CA, USA) for 10 min. After 2 washes with 100 µl FACS buffer, cells were stained with 20 µl of Alexa Fluor® 488 anti-H2A.X Phospho (Ser139) Antibody (613406, Biolegend, San Diego, CA, USA) 1:100 in permeabilization buffer (421002, Biolegend, San Diego, CA, USA) for 30 min. Afterwards, cells were washed twice with 100 µl permeabilization buffer and once with 100 µl FACS buffer and stored in 100 µl FACS buffer at 4 °C. All steps were carried out at 4 °C. Unstained and single stains for all antibodies were included in each run. Alexa Fluor® 488 Mouse IgG1, κ-Isotype Ctrl (FC) Antibody (400132, Biolegend, San Diego, CA, USA) was used as isotype control for Alexa Fluor® 488 anti-H2A.X Phospho (Ser139) Antibody.

The stained and analysed cells were measured as frequent of population (FOP) for all white blood cells, lymphocytes, monocytes and t-cells using Becton Dickinson LSR II flow cytometer (Heidelberg, Germany) and FlowJo version xV0.7 (OR, USA).

### CONSORT 2010 checklist of information to include when reporting a randomised trial*

| Section/Topic | Item No | Checklist item | Reported on page No |
|---|---|---|---|
| **Title and abstract** | | | |
| | 1a | Identification as a randomised trial in the title | 1 |
| | 1b | Structured summary of trial design, methods, results, and conclusions (for specific guidance see CONSORT for abstracts) | 2 |
| **Introduction** | | | |
| Background and objectives | 2a | Scientific background and explanation of rationale | 4 |
| | 2b | Specific objectives or hypotheses | 4 |
| **Methods** | | | |
| Trial design | 3a | Description of trial design (such as parallel, factorial) including allocation ratio | 5 |
| | 3b | Important changes to methods after trial commencement (such as eligibility criteria), with reasons | n.a. |
| Participants | 4a | Eligibility criteria for participants | 5, suppl. |
| | 4b | Settings and locations where the data were collected | 5 |
| Interventions | 5 | The interventions for each group with sufficient details to allow replication, including how and when they were actually administered | 5-6 |
| Outcomes | 6a | Completely defined pre-specified primary and secondary outcome measures, including how and when they were assessed | 6-7 |
| | 6b | Any changes to trial outcomes after the trial commenced, with reasons | n.a. |
| Sample size | 7a | How sample size was determined | 7 |
| | 7b | When applicable, explanation of any interim analyses and stopping guidelines | n.a. |
| Randomisation: | | | |
| Sequence generation | 8a | Method used to generate the random allocation sequence | 5 |
| | 8b | Type of randomisation; details of any restriction (such as blocking and block size) | 5 |
| Allocation concealment mechanism | 9 | Mechanism used to implement the random allocation sequence (such as sequentially numbered containers), describing any steps taken to conceal the sequence until interventions were assigned | 5 |
| Implementation | 10 | Who generated the random allocation sequence, who enrolled participants, and who assigned participants to interventions | 5 |

| | | | |
|---|---|---|---|
| Blinding | 11a | If done, who was blinded after assignment to interventions (for example, participants, care providers, those assessing outcomes) and how | n.a. |
| | 11b | If relevant, description of the similarity of interventions | 5-6 |
| Statistical methods | 12a | Statistical methods used to compare groups for primary and secondary outcomes | 7 |
| | 12b | Methods for additional analyses, such as subgroup analyses and adjusted analyses | 7 |
| **Results** | | | |
| Participant flow (a diagram is strongly recommended) | 13a | For each group, the numbers of participants who were randomly assigned, received intended treatment, and were analysed for the primary outcome | 8, Figure 1 |
| | 13b | For each group, losses and exclusions after randomisation, together with reasons | 8, Figure 1 |
| Recruitment | 14a | Dates defining the periods of recruitment and follow-up | 8 |
| | 14b | Why the trial ended or was stopped | n.a. |
| Baseline data | 15 | A table showing baseline demographic and clinical characteristics for each group | Table 1 |
| Numbers analysed | 16 | For each group, number of participants (denominator) included in each analysis and whether the analysis was by original assigned groups | Table 1, Table 2, Figure 2 |
| Outcomes and estimation | 17a | For each primary and secondary outcome, results for each group, and the estimated effect size and its precision (such as 95% confidence interval) | 8-9, Figure 2, Table 2, suppl. |
| | 17b | For binary outcomes, presentation of both absolute and relative effect sizes is recommended | |
| Ancillary analyses | 18 | Results of any other analyses performed, including subgroup analyses and adjusted analyses, distinguishing pre-specified from exploratory | 8, suppl. Fig.3, suppl. Table S2 |
| Harms | 19 | All important harms or unintended effects in each group (for specific guidance see CONSORT for harms) | n.a. |
| **Discussion** | | | |
| Limitations | 20 | Trial limitations, addressing sources of potential bias, imprecision, and, if relevant, multiplicity of analyses | 13-14 |
| Generalisability | 21 | Generalisability (external validity, applicability) of the trial findings | 14 |
| Interpretation | 22 | Interpretation consistent with results, balancing benefits and harms, and considering other relevant evidence | 12-14 |
| **Other information** | | | |
| Registration | 23 | Registration number and name of trial registry | 2, 5 |
| Protocol | 24 | Where the full trial protocol can be accessed, if available | n.a. |
| Funding | 25 | Sources of funding and other support (such as supply of drugs), role of funders | 15 |

*We strongly recommend reading this statement in conjunction with the CONSORT 2010 Explanation and Elaboration for important clarifications on all the items. If relevant, we also recommend reading CONSORT extensions for cluster randomised trials, non-inferiority and equivalence trials, non-pharmacological treatments, herbal interventions, and pragmatic trials. Additional extensions are forthcoming: for those and for up to date references relevant to this checklist, see www.consort-statement.org.
